# Wastewater-Based Analysis of Antihistamines to Investigate Pollinosis Symptom Burden at Population-Scale

**DOI:** 10.1101/2023.11.29.23299171

**Authors:** Stephan Baumgartner, Michelle Salvisberg, Bernard Clot, Benoît Crouzy, Peter Schmid-Grendelmeier, Heinz Singer, Christoph Ort

**Affiliations:** Swiss Federal Institute of Aquatic Science and Technology, Eawag, Dübendorf, Switzerland; Institute of Civil, Environmental and Geomatic Engineering, ETH Zürich, Zurich, Switzerland; Federal Office of Meteorology and Climatology MeteoSwiss, Payerne, Switzerland; Allergy Unit, Department of Dermatology, University Hospital Zurich, University of Zurich, Zurich, Switzerland; Christine Kühne-Center for Allergy Research and Education, Davos, Switzerland

**Keywords:** Pollinosis, antihistamines, wastewater-based epidemiology, airborne pollen, seasonal allergic rhinoconjuctivitis

## Abstract

**Background:** Pollinosis, or hay fever, is the most prevalent allergic disorder. Assessing the impact of real-world pollen exposure on symptoms remains challenging due to the extensive efforts required at the patient level.

**Objectives:** We explore the potential of wastewater-based epidemiology (WBE) to investigate the impact of exposure to specific pollen taxa on symptoms and to assess self-medication patterns of second-generation antihistamines at a population-scale.

**Methods:** In Zurich (Switzerland), 279 wastewater samples were collected from 2021-2023. Each sample represents a 24-hour period with excreta from approximately 471,000 individuals. Eleven antihistamine markers were analyzed in the samples using liquid chromatography high-resolution mass spectrometry. The relationship between antihistamine loads in wastewater and concentrations of airborne pollen (47 taxa) was investigated using multivariate linear regression analysis.

**Results:** The loads of three second-generation antihistamines in wastewater showed strong day-to-day variation correlating with airborne pollen patterns. About 50% of the annual fexofenadine consumption was linked to acute pollen exposure, 20% to baseline consumption during pollen season, and 30% was independent of pollen. Alder, birch, grasses, hornbeam, plane, and plantain explained most of the variance in consumption (R^2^ = 0.82), with grass pollen alone causing a quarter of the annual consumption. Increased fexofenadine consumption during periods without elevated concentrations of common allergenic pollen suggests the presence of additional triggers for allergy symptoms, potentially yew pollen.

**Conclusions:** Our study demonstrates that WBE can effectively capture substantial day-to-day variation in antihistamine consumption caused by pollen exposure symptoms. As such, WBE is an objective, cost-effective, and questionnaire-independent method for investigating pollinosis at a population-scale.

## 1. Introduction

In recent decades, the prevalence of allergic diseases, particularly in industrialized countries, has been on the rise, presenting a significant health challenge [1,2]. These conditions not only cause discomfort and distress to patients but also impose a substantial economic burden. In Europe, for instance, the management of seasonal allergies incurs annual costs estimated between 50 and 150 billion euros [3]. Among these allergic disorders, pollinosis, also referred to as seasonal allergic rhinoconjunctivitis, or simply hay fever, emerges as the most prevalent, estimated to affect one-fifth of the Swiss population [4].

While pollen exposure serves as the primary trigger for pollinosis, this multifaceted condition is influenced by a diverse range of external factors. These encompass elements such as air pollution, heat, humidity, thunderstorms, and the diversity of pollen allergens [5,6]. Together, these factors not only augment the allergenic potential of pollen, but also influence an individual’s susceptibility to these allergens [7–9]. Additionally, global phenomena like climate change and urbanization are posited to significantly contribute to the shifting epidemiology of seasonal allergies, further amplifying the challenge of understanding and managing these conditions [10–14]. Understanding the intricate interplay of these factors and their impact on pollinosis requires comprehensive monitoring at a population-scale and the triangulation of data with environmental factors. This is challenging with clinical studies focusing on individual patients due to the high level of effort required. To address this gap, alternative approaches such as digital epidemiology and symptom diary apps have been explored [15–17].

However, these methods may be susceptible to limitations in obtaining representative samples, recall bias, and the need for active participations, hindering their full potential. In contrast, wastewater-based epidemiology (WBE) offers objective insights into community-level health without the need for active participation by individuals. WBE has demonstrated its efficacy in monitoring pathogen exposure during the SARS-CoV-2 pandemic and extends its utility to study community-wide consumption of pharmaceuticals, diet and lifestyle products, as well as exposure to industrial chemicals and pesticides [18–20].

Notably, previous applications of WBE have indirectly explored the influence of environmental factors, such as air pollution and heat, on acute asthma and cardiovascular conditions by analyzing pharmaceuticals in wastewater [21,22]. The current study uses WBE to study the population burden of seasonal allergies by investigating the relationship between antihistamine residues excreted to wastewater and airborne pollen concentrations.

## 2. Materials and methods

### 2.1. Chemicals and solutions

Information regarding the chemicals and solutions utilized for the analysis of the eleven antihistamine markers bilastine, cetirizine, desloratadine, 3-hydroxydesloratadine, diphenhydramine, fexofenadine, fexofenadine N-oxide, hydroxyzine, loratadine, meclizine, and rupatadine in wastewater is provided in the supporting information (SI1.2).

### 2.2. Wastewater sample collection and storage

Volume-proportional 24-hour composite samples were collected from the raw influent at the wastewater treatment plant (WWTP) of Zurich using the on-site auto-sampler. Sampling was conducted from February to July 2021 (pollen season) and in December 2021 (pollen off-season) on a daily basis. During the remaining months 2021-2023, samples were collected every 13th day. The wastewater samples were kept at 4°C in the auto-sampler during the 24 hours of sampling and were then transferred into muffled 100mL glass bottles and stored at -20°C until chemical analysis. More information about the WWTP catchment characteristics and the sampling procedure is provided in SI1.1.

### 2.3. Wastewater sample preparation

The wastewater samples were thawed and centrifuged at room temperature at 279.5 x g for 10 minutes (Centrifuge 5427 R, Eppendorf). Subsequently, 600 µL of supernatant were transferred to a new glass vial and spiked with 10 µL of an ethanol-based isotope-labeled internal standard (ISTD) mix containing 9 ISTDs, to a final concentration of 1000 ng/L (SI1.2). A ten-point calibration in Evian mineral water ranging from 10 ng/L to 10 µg/L was prepared from four ethanol-based working standard solutions. For each measurement batch, three independently prepared samples were analyzed to determine the precision of the analytical method. Furthermore, specific samples were spiked with the target analytes at seven concentration levels ranging from 50 ng/L to 5 µg/L to assess analyte recoveries in sample matrix.

For cetirizine the calibration (nine-point ranging from 10 ng/L to 5 µg/L) and the spiked samples (six-point ranging from 50 ng/L to 2.5 µg/L) were prepared independently from ACN:H_2_O (1:1, v/v) -based working solutions analogous to the procedure described above. We additionally measured in-sample stability of target analytes according to experimental conditions outlined in SI3.

### 2.4. LC-HRMS measurements

The wastewater samples were analyzed for the target substances by a large volume direct injection into a reversed-phase chromatograph coupled to high-resolution mass spectrometer (LC-HRMS). The chromatographic system consisted of a PAL auto-sampler (CTC Analytics, Switzerland) and a Dionex UltiMate3000 RS pump (Thermo Fisher Scientific, USA). A sample volume of 100 µL was injected on a reversed-phase C18 column (Atlantis® T3 3 µm, 3.0 x 150 mm; Waters). The chromatographic separation was performed at a flow rate of 300 µL/min with a mobile phase gradient, starting with 100% of eluent A (water and 0.1% v/v formic acid). After 1.5 minutes, eluent B (methanol and 0.1% v/v formic acid) was increased for 17 minutes to 95%, held for 10 minutes, and lowered again to the starting conditions, followed by a 4-minute re-equilibration phase. The samples were measured on a hybrid quadrupole-orbitrap high-resolution mass spectrometer (Orbitrap ExplorisTM 240 from Thermo Fisher Scientific) in positive ionization mode (ESI, 3.5 kV). Full-scans were recorded at a resolution (R) 120 000 (at m/z 200) followed by five data-dependent MS/MS experiments (R = 30 000 at m/z 200). Fragmentation of target ions was triggered based on an inclusion mass list and performed using higher energy collision-induced dissociation (HCD) with stepped collision energies of 15, 45, and 90%.

The HRMS measurements were evaluated with TraceFinder 5.1 (Thermo Fisher, USA). Target analytes were quantified from the extracted ion chromatogram of the MS full-scan based on the area ratio of the reference standard (STD) and the corresponding ISTD of the analyte.

Detailed information on the LC-HRMS settings and measurement quality control is presented in SI1.

### 2.5. Daily population loads in wastewater

Substance-specific daily population-normalized wastewater loads (*DPL*, mgd^-1^1000p^-1^) were determined by multiplying the concentrations measured in the wastewater samples (*C*_*sample*_, µgL^-1^) with the daily wastewater volume (*Flow*_*WTP*_, m^3^d^-1^) divided by the estimated population in the catchment (*Pop*_*WTP*_, 1000p), as described in Equation 1.

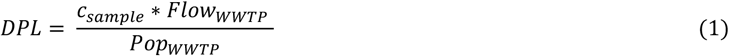

### 2.6. Pollen measurements

Pollen data from the operational measurements of the Swiss pollen network at the Federal Institute of Meteorology and Climatology (MeteoSwiss), were utilized in this study. The quantification of airborne pollen was conducted in alignment with standardized procedures (EN 16868:2019-09) [23].

The characteristics of the pollen measurement station are detailed in SI1.5.

### 2.7. Model to describe relationship between airborne pollen and fexofenadine loads in wastewater

We used a linear model to characterize the relationship between airborne pollen exposure and antihistamine residues in wastewater at the population scale (Equation 2). In this descriptive model, the fexofenadine load (*DPL*_*t*_; mgd^-1^1000p^-1^) is a function of a baseline value (*b*_*t*_; mg d^-1^ 1000p^-1^), the airborne concentrations of the *n* allergenic pollen taxa (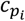; pollen m^-3^) and weighting factors 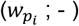.

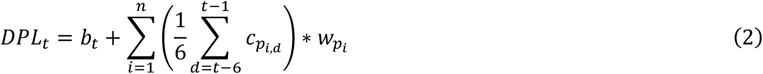

We observed fexofenadine loads during periods of negligible pollen exposure, mainly but not exclusively in winter months. Therefore, a baseline was needed to describe presumably pollen-independent consumption. We defined a day as a baseline point if no allergenic pollen taxa exceeded the lowest load classification (SI1.6) in the nine preceding days. The baseline for the remaining days was derived from linear interpolation between baseline points. The baseline values from 2021 were also used for 2022 and 2023, as the lower temporal coverage of sample analysis in these years did not allow for the determination of an independent baseline. The fexofenadine loads exceeding the baseline were described by a pollen-dependent term. The pollen concentrations 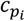 *were t*ransformed (smoothing and applying a temporal offset) by using mean values from the six previous days (SI2.4.1).

Since we observed indications of a linear relationship between transformed pollen concentrations and residual fexofenadine loads (*DPL - b*) (SI2.4.2), and assume additive contributions of pollen, we used Non-Negative Least Squares (NNLS) and Ordinary Least Squares (OLS) regression to determine the taxa-specific coefficients 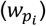. Diagnostic information for the NNLS and OLS regressions is provided in SI2.4.3.

All analyses were conducted in Python 3.10 using the scipy 1.13.0 package for NNLS optimization and statsmodels 0.14.0 package for OLS regression. Access to code and computational environments used for evaluation is provided in the data availability section.

## 3. Results

### 3.1. Antihistamine dynamics in wastewater

Out of the eleven antihistamine markers (bilastine, cetirizine, desloratadine, 3-hydroxydesloratadine, diphenhydramine, fexofenadine, fexofenadine N-oxide, hydroxyzine, loratadine, meclizine, and rupatadine) analyzed in the wastewater samples, four were detected in measurable quantities in the majority of samples: bilastine, cetirizine, fexofenadine, and diphenhydramine (SI2.2). The absence of quantifiable amounts for the other compounds can be attributed to several factors, including: i) low prescribed doses and excretion rates (as for loratadine, desloratadine, and 3-hydroxydesloratadine), ii) low stability in the wastewater matrix (seen in the case of meclizine and fexofenadine N-oxide), iii) the lack of registered products in Switzerland (rupatadine), and iv) relatively poor ionization properties (notably desloratadine and meclizine) leading to higher limits of quantification (LOQ) using the current analytical method (SI2.1 and SI2.2).

We excluded samples collected between 13.7.2021 and 18.7.2021 due to heavy rain and potential discharges of untreated wastewater through combined sewer overflows, leading to underestimation of daily loads. Additionally, outliers for fexofenadine on 18.4.2021, cetirizine on 23.3.2021 and 15.2.2021, and diphenhydramine on 10.6.2021 and 11.2.2021 were excluded due to unusually high values and suspicion of disposal of unconsumed medication in the wastewater. The wastewater loads of second-generation antihistamines, including fexofenadine, bilastine, and cetirizine, demonstrated pronounced seasonal patterns that exhibited a strong correlation among them (Figure 1).

**Figure 1.**
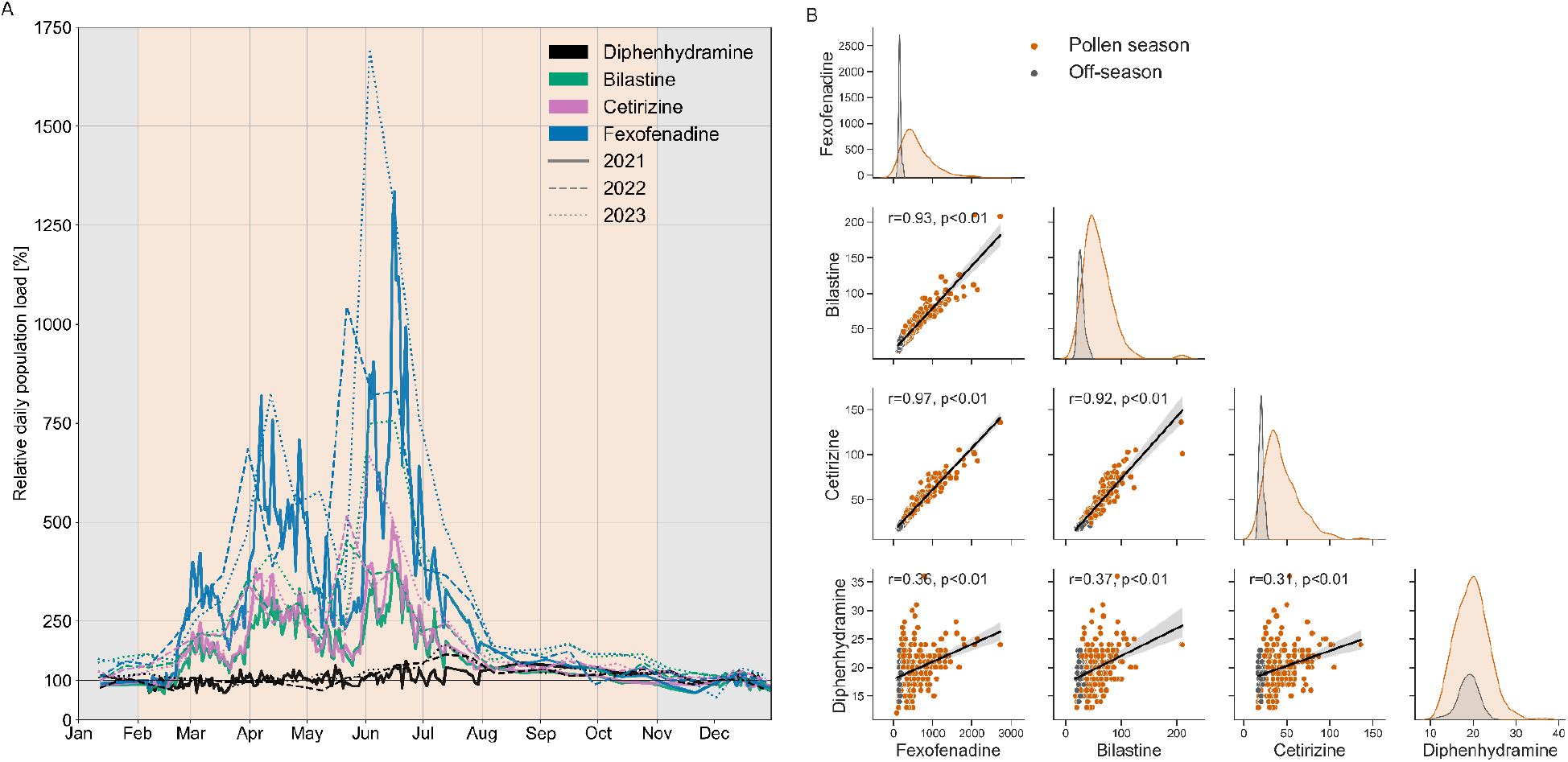
Temporal patterns of antihistamine wastewater populations loads in Zurich during 2021, 2022 and 2023. **A** Antihistamine population loads relative to the average off-season values of January, November, and December (grey). **B** Correlation of wastewater population loads [mg/d/1000p] with Pearson’s r and associated p-values.

Outside pollen season, in late summer, fall and winter, the wastewater loads remained consistently low (Figure 1A). However, distinct periods in spring and summer displayed a significant surge, with fexofenadine loads ten-fold or higher compared to winter levels.

### 3.2. Airborne pollen patterns and fexofenadine wastewater loads

Fexofenadine was chosen as the representative compound for second-generation antihistamines in the descriptive modeling of the relationship between pollen exposure and antihistamine consumption, due to its detectability above the LOQ in all samples and its high stability (SI2.2 and SI3.3). The observed high correlation among the loads of second-generation antihistamines further justifies the focus of this analysis (Figure 1B).

Out of the 47 pollen taxa analyzed in this study, we paid special attention to i) 14 allergenic taxa: alder, ash, beech, birch, grasses, hazel, hornbeam, mugwort, oak, plane, plantain, ragweed, sorrel and sweet chestnut, since these make up the Swiss pollen burden monitoring and ii) the 5 taxa: cypress family, olive family, nettle family, pine and yew for which indications of allergenicity have been reported previously.

The pollen data of allergenic taxa that were found to contribute significantly (p < 0.01) to fexofenadine consumption in OLS (Table SI11) are displayed in Figure 2, while the remaining data of the total 47 taxa and miscellaneous pollen are presented in SI2.3.

**Figure 2.**
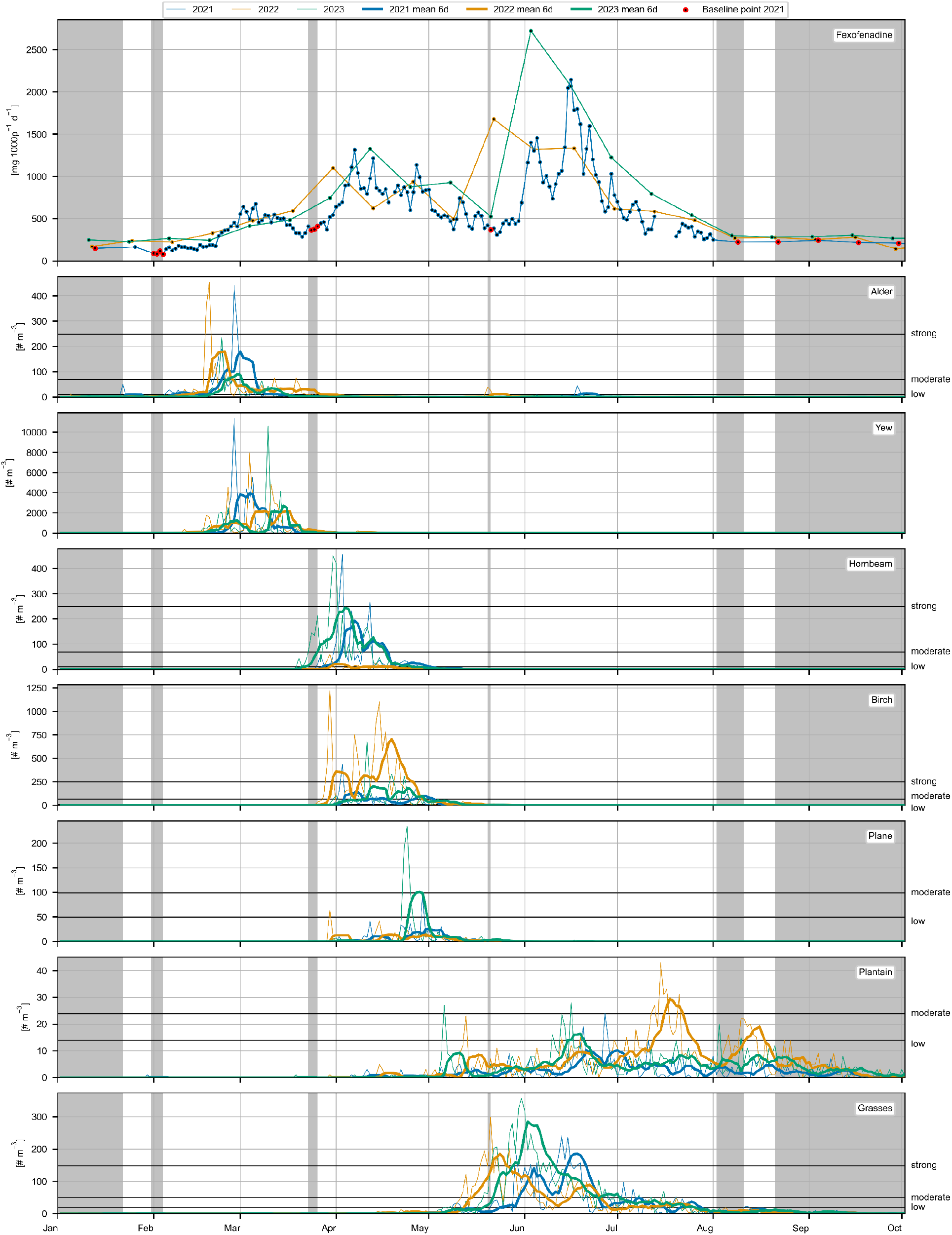
Concentrations of allergenic airborne pollen (number of pollen grains/m^3^) and fexofenadine wastewater loads (mg/day/1000 people) in Zurich during 2021, 2022, and 2023. Upper limits of pollen taxa-specific load classes are indicated by horizontal lines. Grey shading indicates days in 2021 when no pollen taxa exceeded the lowest load class in the previous nine days, with red circles highlighting the corresponding fexofenadine loads as baseline points. Thin lines show actual observed pollen concentrations, while thick lines represent the mean pollen values of the previous six days used for modeling.

In the years 2021 to 2023 the annual pattern of allergenic pollen roughly divides into three periods delineated by phases of low pollen concentrations. All three pollen periods coincide with elevated wastewater loads of fexofenadine and correspondingly low loads during phases of low pollen burden (Figure 2).

During the first period, spanning from January to the end of March, the early pollen of hazel and alder dominate among the allergenic taxa. Additionally, we observed exceptionally high concentrations of yew pollen during this time, surpassing the maximum concentrations of the other pollens by an order of magnitude. The second period, lasting from the end of March to mid-May, is characterized by elevated concentrations of the allergenic pollen of ash, hornbeam, birch, oak, plane and beech (Figure 2 and Figure SI3). In the last period, from late May to the end of July, grass pollen dominate, coinciding with low pollen concentrations of sweet chestnut and plantain (Figure 2 and Figure SI4). Relatively low pollen concentrations were observed for the allergenic taxa of mugwort (maximum 8 pollen/m^3^), ragweed (maximum 11 pollen/m^3^), and sorrel (maximum 16 pollen/m^3^).

### 3.3. Multivariate linear regression analyses

In the descriptive model, the temporal pattern of fexofenadine wastewater loads is described by two main components: i) baseline consumption, which is independent of specific pollen concentrations, and ii) consumption associated with exposure to specific pollen taxa (Equation 2).

In Figure 2, we further divided the baseline into two subcategories: i) *baseline off-season*, which refers to mean fexofenadine consumption during winter months November to January when allergenic pollen are typically absent, and ii) *baseline pollen season*, which refers to increased background consumption during spring and summer months that persists despite minimal pollen concentrations over a nine-day period. The remainder above the two baselines, we refer to as *residual fexofenadine loads*.

To analyze the relationship between fexofenadine residual loads and pollen concentrations of specific taxa, we found that transforming pollen concentrations using the mean of the preceding six days (smoothing and applying a temporal offset) resulted in the highest descriptive power (R^2^ = 0.84). Therefore, this transformation was applied to the pollen concentrations for all descriptive modeling. Due to the temporal synchronicity in the pollen patterns of different taxa (Figure SI17), we used several combinations of pollen taxa as explanatory variables in NNLS and OLS to determine the significant contributors.

In NNLS contributions of individual pollen taxa are constrained to be positive, which is the intuitive approach for determining the pollen-specific coefficients. However, we additionally employed OLS in our analysis to obtain further statistical insights. Results from alternative pollen concentration transformations and diagnostic information for NNLS and OLS are provided in SI2.4.

In the various modeling scenarios, whether considering 14 or 19 pollen taxa in NNLS or OLS, we consistently identified five pollen taxa as major contributors to fexofenadine consumption (Figure1): grasses, birch, hornbeam, plane, and plantain. This finding is supported by p-values below 0.01 in the OLS results for these pollen taxa, and the coefficients from NNLS falling within the confidence intervals determined in the OLS analysis (SI2.4.3). Even when considering all 47 pollen taxa and miscellaneous pollen as explanatory variables in NNLS, the main contributions were still estimated to originate from these five pollen taxa (Figure SI19). For observed fexofenadine consumption from February to the end of March, the modeling did not yield consistent results. Considering only the 14 allergenic pollen taxa, alder was a significant (p<0.01) contributor, but an unexplained two-week period in March remained (Figure 3A). When including all 19 allergenic taxa, yew significantly contributed (p<0.01) and, while showing a partly synchronized pattern with alder, explained most of the elevated fexofenadine loads in March (Figure 3B). Since all terms of the model (Equation 2) are additive, the colored areas in Figure 3 can directly be interpreted as estimated contributions of the individual variables.

**Figure 3.**
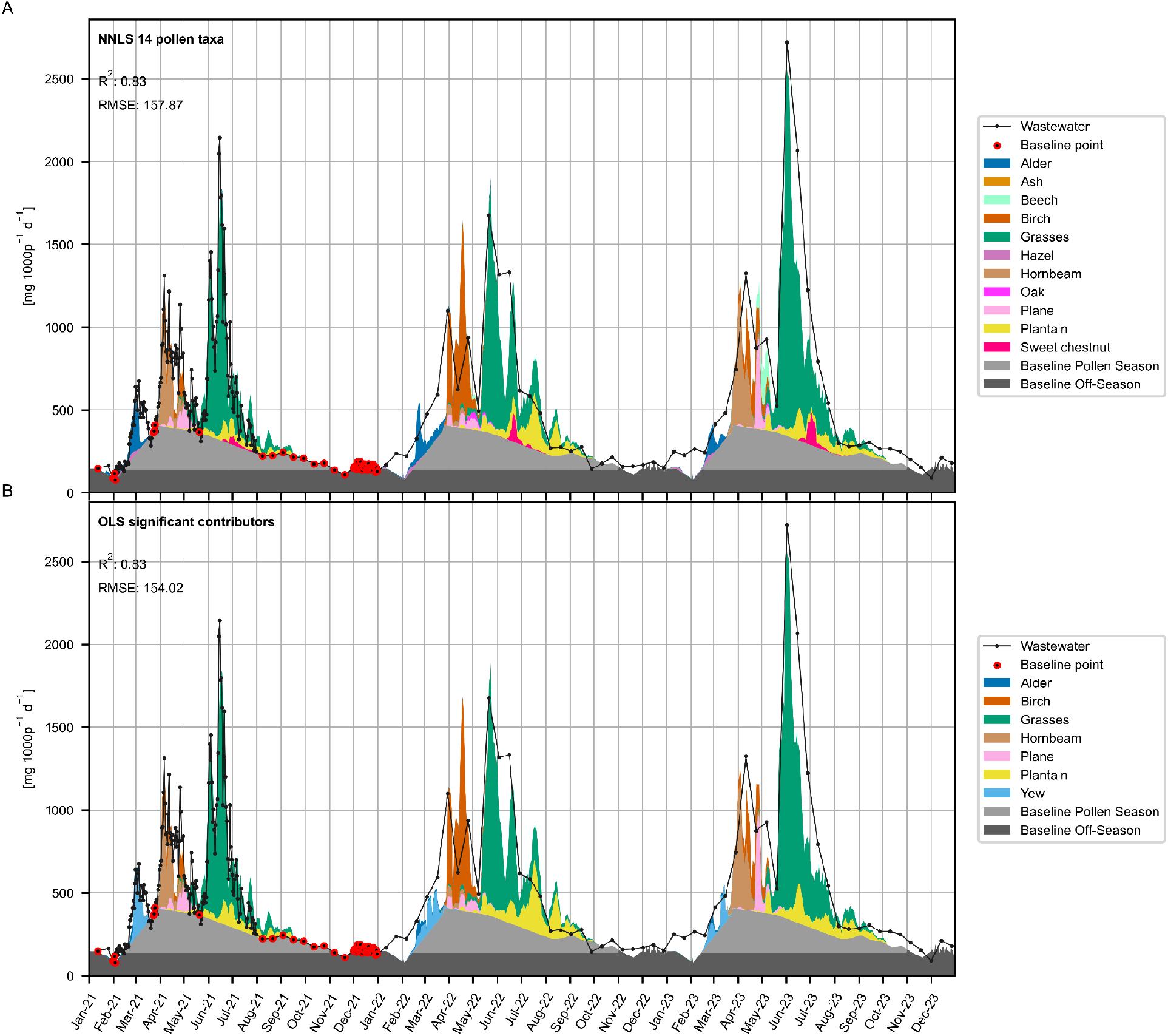
Observed (black line and dots) and predicted (grey and colored shading) fexofenadine wastewater loads in Zurich from 2021 to 2023. The figures display stacked estimated baseline contributions and predicted fexofenadine residual loads based on different models: **A** NNLS using 14 allergenic pollen taxa as explanatory variables and **B** OLS using only pollen taxa identified as significant (p<0.01) positive contributors from the OLS models with 14 or 19 allergenic pollen taxa as explanatory variables. The colored areas represent the estimated, pollen-taxa specific contributions calculated by multiplying the coefficient with the transformed pollen concentration of the respective pollen taxa and day. Baseline is determined by interpolation between fexofenadine loads on days in 2021 when no pollen taxa exceeded the lowest load class in the previous nine days. This baseline is further divided into *baseline pollen season*, which results from subtracting the *baseline off-season* (mean of fexofenadine loads in January, November and December 2021) values from the total baseline value of the respective day. The coefficient of determination (R^2^) and the root mean square error (RMSE) refer to the prediction of the pollen-dependent fexofenadine residual loads (baseline subtracted).

For additional pollen taxa, the NNLS coefficients were estimated to be greater than zero, but they did not result in positive significant (p<0.01) OLS coefficients. The addition of these taxa further led to only a small increase in the explanatory power of the model (R^2^ = 0.85 for NNLS with 47 pollen taxa and miscellaneous pollen). The increased fexofenadine loads measured over several days in mid-May, in the absence of allergenic pollen, could not be explained by any of the 47 pollen taxa or miscellaneous pollen (Figure SI19).

## 4. Discussion

The strong correlation among the three second-generation antihistamines fexofenadine, bilastine and cetirizine, observed in our study suggests similar consumption behaviors. As anticipated, the control substance and first generation antihistamine diphenhydramine, primarily prescribed as a sedative, exhibited consistent loads throughout the year.

Given the prominent use of second-generation antihistamines for treating allergic rhinitis, the observed general seasonal pattern in wastewater was expected. However, the poorly understood self-medication behaviors with these primarily over-the-counter drugs [24,25], limited our knowledge about consumption variations. Our detailed analysis reveals that approximately 30 percent of the annual fexofenadine load is consumed independently of pollen season (Baseline Off-Season). This is likely attributable to the treatment of non-seasonal allergies or non-allergic conditions such as mastocytosis. The remaining 70 percent of fexofenadine consumption is seasonally driven. Notably, around 20 percent of the consumption occurs in a manner not directly linked to acute pollen exposure, as evidenced by its persistence even after 9 consecutive days of negligible pollen concentrations (Baseline Pollen Season). We attribute this to long-term prophylactic treatment for pollinosis symptoms or the treatment of pollen-independent seasonal allergies.

The remaining 50 percent of fexofenadine consumption is associated with the acute treatment of symptoms triggered by pollen exposure. The temporal off-set between pollen exposure and observations fexofenadine loads in wastewater can be explained by the time required for realizing symptoms, consuming drugs, drug elimination, excretion, and in-sewer transport to the sampling point at the influent of the wastewater treatment plant, overall estimated at 1-2 days (Table SI 1 and Table SI 6). Additionally, ongoing symptoms or preventive drug use by allergy sufferers post-exposure could contribute to the delay.

Moreover, the fexofenadine loads exhibited substantial day-to-day variations, driven by the presence or absence of airborne pollen. This was particularly evident in the daily measurements during the grass pollen period in June 2021, when loads trippled within three days and then halved over five days. This indicates that major consumption of second-generation antihistamines in the catchment of Zurich (approximately 471`000 people) is linked to acute symptoms. This means individuals consume the drug based on exposure and stop relatively quickly once the allergen is no longer present, rather than using it continuously over several days to weeks in anticipation of future allergen exposure. Additionally, this linear relationship between exposure and antihistamine consumption scales well, as observed during the very high grass pollen burden in 2023.

This highlights the usefulness of WBE, showing that wastewater data – generally, but in our case particularly daily values – is more nuanced than data obtained from traditional approaches. The latter often rely on large spatial or temporal aggregation to assess symptoms at a population scale, e.g. digital epidemiology (e.g. Google searches, tweets) [16,26,27] or sales data [28]. Unlike sales data, wastewater monitoring directly assesses consumption, avoiding biases from temporal or spatial differences between purchasing and consumption. Furthermore, it does not rely on active participation, as is the case for symptom diary apps [29].

Our data suggests that grasses are estimated to be the largest contributors to fexofenadine consumption, accounting for roughly 25 percent of the total annual load. In comparison, the birch-homologous group, including birch, alder, hazel, oak, and hornbeam [30], contributed to approximately 10 percent of the annual fexofenadine load. These results align with high sensitization reported for grass and birch allergens in Swiss students [31]. However, the pollen taxa-specific burdens can vary greatly between years, as observed particularly for birch, hornbeam, and grass pollen.

The observed preliminary indications of the potential allergenicity of yew pollen at a population scale necessitate further immunological assessment. To date, yew pollen has been largely disregarded as an allergen due to its coniferous nature, despite occasional case studies suggesting otherwise [32,33]. Moreover, reported cross-reactivity between yew pollen allergy and the semi-synthetic taxane cancer drug docetaxel [34,35], combined with the lack of research on potential cross-reactivity with common tree allergens and the absence of yew-specific diagnostic tools, underscores the need for further investigation. It is particularly noteworthy that our study location, Zurich, is home to one of the largest yew tree populations in Europe [36,37]. The measured pollen concentrations were considerably high, exceeding 10,000 pollen/m^3^.Additional pollen types with potential allergenicity, such as the cypress family, olive family, nettle family, and pine, currently do not seem to have allergenic potential at the population level, but this may change with progression of climate change [12,38].

Subsequently, we discuss some open points. First, we assume that second-generation antihistamines are consumed based on symptoms experienced; however, this requires that individuals correctly interpret their symptoms as allergic rhinitis, which is not necessarily the case. We suspect this could include, for example, mistaking allergic rhinitis symptoms for those of a common cold, or vice versa. This can lead to bias and may also explain the unexpectedly low antihistamine consumption at the beginning of the year, despite the presence of hazel pollen. Additionally, the scope of this study necessitated specific simplifications in modeling the relationship between pollen exposure and antihistamine consumption: e.g. determining baseline levels, transforming pollen concentrations, and discerning pollen-specific contributions. While the 13-day sampling over two years was useful to determine boundaries in the modeling, future studies should focus on daily wastewater data to capture important day-to-day variance in symptoms due to acute pollen exposure as seen in our data in 2021. Inclusion of additional contextual factors like short-acting β2 adrenergic receptor agonists (such as salbutamol) [39] and endogenous histamine urine metabolites (for instance, 1,4-methylimidazole acetic acid) [40] could provide further insights into the relationship with asthmatic symptoms and histamine-mediated conditions. However, the quantification of histamine metabolites presents notable analytical challenges [41], and their potential as biomarkers is presumably limited to systemic mastocytosis rather than allergen exposure [42].

Concluding, our study shows that WBE can effectively capture the significant day-to-day variance in symptoms experienced due to pollen exposure, making it a promising, objective, cost-effective and questionnaire-independent method for studying the effects of pollen allergy at a population scale. This opens doors for future WBE applications for discerning pivotal factors influencing symptom severity and recognizing overlooked allergenic triggers, as exemplified by our observations on yew pollen. These factors also include the impact of environmental influences such as air pollution, weather conditions, global warming-related changes in blooming patterns and plant composition as well as the advent of invasive species like ragweed on allergic conditions. Advancements in automated pollen measurements, potentially providing improved spatial and temporal pollen information, along with the global trends towards the institutionalization of wastewater monitoring, offer a promising foundation for future studies in this direction [43–46].

## Data Availability

The full dataset, along with the associated analysis and visualization scripts, will be made available to the public through the Eawag Research Data Institutional Collection (ERIC-open).

## Abbreviations used

ACN: Acetonitrile
HCD: higher energy collision-induced dissociation
ISTD: isotope-labeled internal standard
LC-HRMS: liquid chromatography high-resolution mass spectrometry
LOQ: limit of quantification
NNLS: non-negative least squares
OLS: ordinary least squares
WBE: wastewater-based epidemiology
WWTP: wastewater treatment plant

## Author contributions

Conceptualization: SB, HS, CO, PSG; Methodology: SB, MS, HS, BCL, BCR; Measurements: SB, MS; Resources: SB, CO, HS, PSG, BCL, BCR; Original Draft: SB; Review & Editing: SB, PSG, CO, HS; Final submission: all authors

## Acknowledgments

The authors would like to thank the personnel of the WWTP Zurich, with particular acknowledgment to Rey Eyer for his efforts in sampling. We also express our sincere appreciation to Philipp Longrée for his support with the LC-HRMS analysis, and to Andreas Scheidegger and Patrick Schmidhalter for their expertise and assistance in statistical analysis.

AI-assisted technology, ChatGPT, was utilized exclusively for language improvement in the manuscript. The authors take full responsibility for the content of the publication.

This research received funding from the Swiss Federal Office of Public Health (FOPH, contract number: [142003899/321-446/1]). Eawag further supported the study with additional funds to enable daily sample analyses.

