## Supporting Information for "Wastewater-Based Analysis of Antihistamines to Investigate Pollinosis Symptom Burden at Population-Scale"

### Contents

SI 1. Materials and Methods ... 3
SI 1.1 Wastewater treatment plant (WWTP) characteristics and wastewater sampling procedure ... 3
SI 1.2 Chemicals and Solutions ... 4
SI 1.3 Analytical Instrumentation and Method ... 5
SI 1.4 Quantification Method and Quality Control ... 6
SI 1.4.1 Quantification ... 6
SI 1.4.2 Matrix effects ... 6
SI 1.4.3 Limit of quantification ... 6
SI 1.4.4 Accuracy ... 6
SI 1.4.5 Precision ... 7
SI 1.5 Pollen measurement station characteristics ... 7
SI 1.6 Pollen load classes ... 7
SI 2. Results and Discussion ... 8
SI 2.1 Antihistamine substance properties ... 8
SI 2.2 LCMS Quantification ... 9
SI 2.3 Airborne pollen patterns and fexofenadine wastewater patterns ... 11
SI 2.4 Pollen antihistamine model ... 17
SI 2.4.1 Pollen concentration transformation ... 17
SI 2.4.2 Linearity of pollen antihistamine correlation ... 19
SI 2.4.3 Multivariate linear least squares analysis. ... 19
SI 3. In-sample stability of antihistamines ... 32
SI 3.1 Sample preparation and analytical procedure ... 32
SI 3.2 Data analysis ... 33
SI 3.3 Stability results ... 33
SI 4. References ... 36

### SI 1. Materials and Methods

#### SI 1.1 Wastewater treatment plant (WWTP) characteristics and wastewater sampling procedure

Table SI 1: Info on WWTP and on the wastewater sampling procedure

| Characteristic | Value |
|---|---|
| City | Zurich |
| WWTP name | ARA Werdhölzli |
| Date | 31.5.2023 |
| Catchment population | 471`275 |
| Population estimation source | census |
| N pump stations | 38 |
| Maximal sewer distance [km] | 15 |
| Mean sewer residence time [h] | 1 |
| Maximal sewer residence time [h] | 3 |
| Flow meter location | After screen, oil and sand trap |
| Flow meter type | Venturi |
| Dry weather volume [$m^3$/d] | 143`655 |
| Minimal flow rate at dry weather [L/s] | 750 |
| Maximal flow rate at dry weather [L/s] | 3000 |
| Maximal flow rate at rainy weather [L/s] | 6000 |
| Sampling location | After fine screen |
| Sampling device | WaterSam WS316 MS3 |
| Sampling storage temp. [°C] | 4 |
| Sampling container material | Polyethylene (PE) |
| Sampling mode | volume-proportional |
| Sampling interval [$m^3$] | 800 |
| Sampling time [h] | 24 |
| Time until samples are frozen [h] | 7 |
| Sample storage temp. [°C] | -20 |

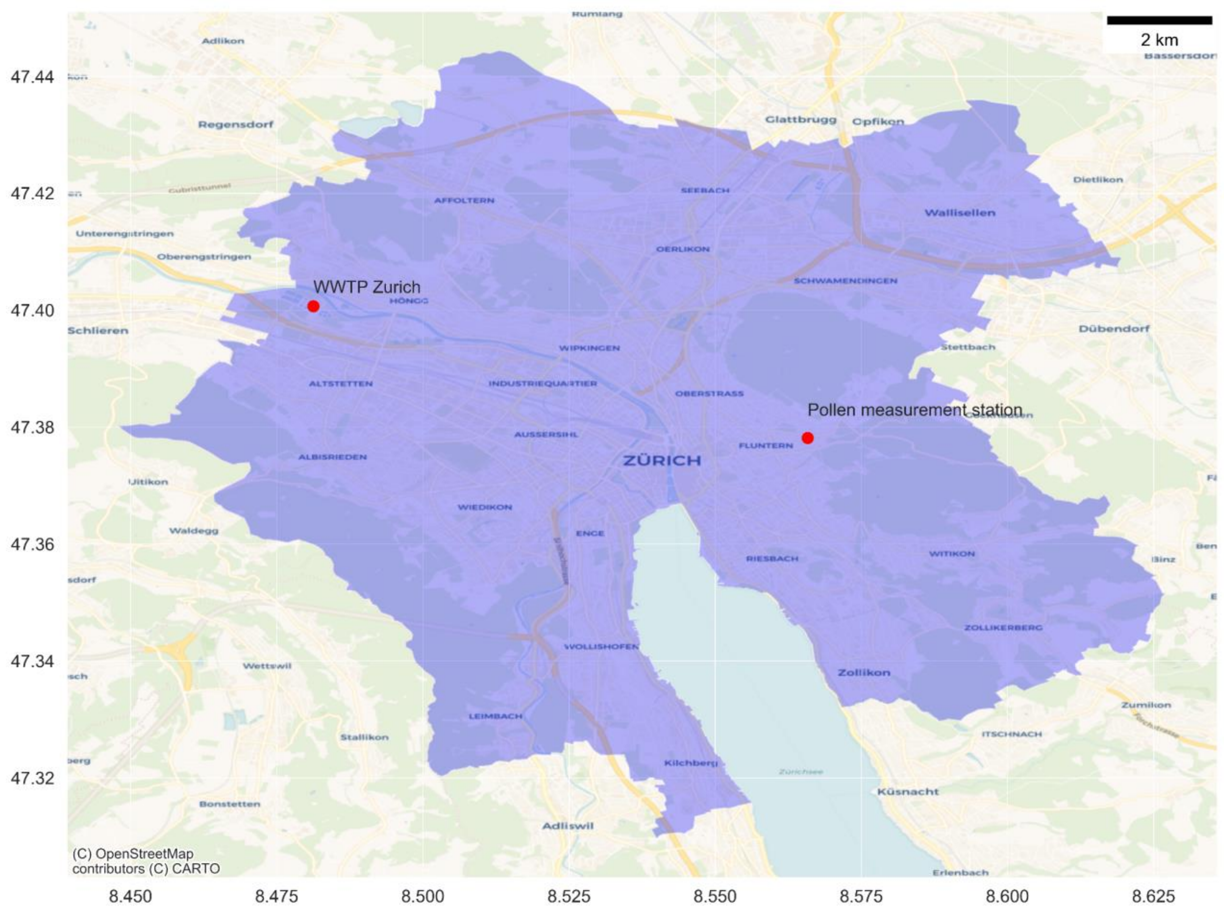


Figure SI 1: Catchment of w astewater treatment plant (WWTP Zurich). The catchment area is shaded in blue. The locations of the WWTP and pollen measurement station are marked w ith red dots.

#### SI 1.2 Chemicals and Solutions

Organic solvents w ere of HPLC grade purity (> 98%) and supplied by Merck (ethanol) or Fisher Chemical (methanol and acetonitr ile). Ultrapure w ater was obtained from a laboratory purification system (Arium Pro, Sartorius). Concentrated formic acid (>98%) w as supplied by Merck.

Stock solutions of the individual analytes (STDs & ISTDs) w ere prepared at a concentration of 1 g/L in ethanol, methanol, or in acetonitrile:w ater (1:1) as described in Table SI 2. Individual stock solutions w ere combined into w orking mix solutions of 500, 60, 6, and 0.6 µg/L in EtOH. For cetirizine independent w orking solutions in ACN:$H_2O$ (1:1, v/v) at the same concentrations were prepared.
The isotope-labelled standard mix w as prepared in ethanol w ith a concentration of 60ug/L. All solutions w ere stored at -20°C except for cetirizine solutions, w hich were stored at 4°C.

Table SI 2: Analytical reference standards (STD) and corresponding isotope-labelled standards (ISTD)

| Substance | CAS-No | Molecular formula | Manufacturer | Solvent | Corresponding ISTD |
|---|---|---|---|---|---|
| 3-Hydroxydesloratadine | 119410-08-1 | C19H19ClN2O | TRC-Canada | EtOH | Desloratadine-D4 |
| Bilastine | 202189-78-4 | C28H37N3O3 | Cayman Chemical | MeOH | Bilastine-D6 |
| Cetirizine | 83881-51-0 | C21H25ClN2O3 | Lipomed AG | ACN:H2O (1:1, v/v) | Cetirizin-D8 |
| Desloratadine | 100643-71-8 | C19H19N2Cl1 | TRC-Canada | EtOH | Desloratadine-D4 |
| Diphenhydramine | 58-73-1 | C17H21NO | Cerilliant | MeOH | Diphenhydramine-D3 |
| Fexofenadine | 83799-24-0 | C32H39N1O4 | TCI Europe | MeOH | Fexofenadine-D6 |
| Fexofenadine N-oxide | 1422515-52-3 | C32H39NO5 | LGC Standards | MeOH | Fexofenadine-D6 |
| Hydroxyzine | 68-88-2 | C21H27ClN2O2 | Lipomed AG | MeOH | Hydroxyzine-D8 |
| Loratadine | 79794-75-5 | C22H23ClN2O2 | Sigma-Aldrich | EtOH | Oxazepam-D5 |
| Meclizine | 569-65-3 | C25H27N2Cl1 | LGC Standards | EtOH | Meclizine-D8 |
| Rupatadine | 158876-82-5 | C26H26N3Cl1 | Cayman Chemical | MeOH | Rupatadine-D4 |
| Fexofenadine-D6 | 548783-71-7 | C32H33[2]H6N1O4 | TRC-Canada | MeOH | - |
| Meclizine-D8 | - | C25H19[2]H8N2Cl1 | TRC-Canada | MeOH | - |
| Cetirizin-D8 | 774596-22-4 | C21H17[2]H8N2O3Cl1 | TRC-Canada | EtOH:H2O (1:1, v/v) | - |
| Diphenhydramine-D3 | 170082-18-5 | C17H18[2]H3N1O1 | Supelco | MeOH | - |
| Desloratadine-D4 | 381727-29-3 | C19H15[2]H4N2Cl1 | TRC-Canada | EtOH | - |
| Hydroxyzine-D8 | - | C21H19[2]H8N2O2Cl1 | Lipomed AG | MeOH | - |
| Bilastine-D6 | - | C28H31[2]H6N3O3 | TRC-Canada | MeOH | - |
| Rupatadine-D4 | - | C26H22[2]H4N3Cl1 | TRC-Canada | MeOH | - |
| Oxazepam-D5 | 65854-78-6 | C15H6[2]H5N2O2Cl1 | Lipomed AG | ACN | - |

#### SI 1.3 Analytical Instrumentation and Method

Table SI 3: Parameter settings used for HRMS/MS measurements with Exploris$^{TM}$ 240 mass spectrometer

| **Parameter** | **Value** |
|---|---|
| Ionization mode | positive |
| Spray voltage [kV] | 3.5 |
| Ion transfer tube temperature [°C] | 320 |
| Sheet gas flow (nitrogen) [AU] | 40 |
| Auxiliary gas flow (nitrogen) [AU] | 10 |
| Sweep gas flow [AU] | 0 |
| RF Lens [%] | 60 |
| Spectrum data type | Profile |
| Maximum injection time MS1 [ms] | 100 |
| Scan range MS1 [m/z] | 100 - 1000 |
| Resolution MS1 | 120000 |
| Internal calibration | Yes (Run Start EASY-IC) |
| Data-dependent mode | Number of scans |
| Data-dependent trigger | Ions of mass list |
| Isolation window [m/z] | 1 |
| Resolution MS/MS | 30000 |
| Collision energy mode | Stepped |
| Collision energy type | Normalized |
| HCD collision energies [%] | 15, 45, 90 |

AU: arbitrary units
ms: milliseconds
m/z: mass to charge ratio

#### SI 1.4 Quantification Method and Quality Control

##### SI 1.4.1 Quantification

Target analytes were quantified based on the area ratio of the analyte and its selected isotope-labelled internal standard (ISTD) using Tracefinder 5.1 software. Structurally identical ISTDs were available for most analytes, except for the transformation products 3-hydroxydesloratadine, fexofenadine N-oxide, and loratadine. The target peaks were extracted with a mass tolerance of 5 ppm, and only peaks with at least five data points and a signal-to-noise ratio greater than 10 were considered for quantification. Extracted peaks were verified as target analytes by comparing the MS/MS spectra, retention time, and isotopic pattern to those of the corresponding reference standards. The ten-point calibration in Evian mineral water included the following concentration levels: 10, 25, 50, 100, 250, 500, 1000, 2500, 5000, 10000 ng/L.
A quadratic calibration curve was employed for quantification of loratadine, desloratadine, and 3-hydroxydesloratadine, while a linear calibration curve was used for all other substances. Regressions were weighted with 1/x.
All quality control parameters of the LC-HRMS measurements that are described below were determined for each measurement batch independently.

##### SI 1.4.2 Matrix effects

The impact of matrix effects during analysis was evaluated by determining the absolute recoveries specific to each analyte. For target analytes with structurally identical ISTD, the absolute recovery is described as the ratio of the ISTD area in the matrix samples ($ISTD\ Area_{Matrix}$) to their average area in the calibration series ($Average\ ISTD\ Area_{Calibration}$), as described in equation SI 1:

$$Absolute\ Recovery = \frac{ISTD\ Area_{Matrix}}{Average\ ISTD\ Area_{Calibration}} \qquad (SI\ 1)$$

If a structurally identical ISTD was unavailable, the absolute recovery was calculated based on the difference of the target peak area in spiked samples ($STD\ Area_{spiked\ sample}$) and the area in the unspiked sample ($STD\ Area_{unspiked\ sample}$) and set into relation with the area in the respective calibration standard ($STD\ Area_{Calibration}$) (Equation SI 2).

$$Absolute\ recovery_{non-identical\ ISTD} = \frac{STD\ Area_{spiked\ sample} - STD\ Area_{unspiked\ sample}}{STD\ Area_{corresponding\ calibration\ standard}} \qquad (SI\ 2)$$

##### SI 1.4.3 Limit of quantification

The limit of quantification (LOQ) was determined based on the lowest concentration in the wastewater matrix, at which the quantification criteria (i. signal-to-noise ratio larger than 10 and ii. at least five data points) was met. If such an LOQ in the wastewater matrix was not determinable, the LOQ was determined based on the lowest quantified calibration level multiplied by the inverse of the absolute recovery (Equation SI 3).

$$LOQ_{matrix}\ [ngL^{-1}] = \frac{LOQ_{solvent}}{Absolute\ recovery} \qquad (SI\ 3)$$

Additionally, if the target analyte was detected in the matrix blanks (Evian mineral water spiked with ISTD and processed as the samples) an LOQ based on the equation below, with sd being the standard deviation, was calculated (Equation SI 4).

$$LOQ_{matrix\ blank}\ [ngL^{-1}] = Average\ C_{matrix\ blank} + 10 * sd(C_{matrix\ blanks}) \qquad (SI\ 4)$$

If multiple LOQ values were obtained the higher value was considered.

##### SI 1.4.4 Accuracy

The accuracy of the measurements was assessed by determining the relative recovery in wastewater samples that were deliberately spiked with specific concentrations as described in the equation below. In each measurement batch, a wastewater sample was spiked with the target analytes at the following concentrations; 50, 100, 250, 500, 1000, 2500, and 5000 ng/L. Cetirizine was spiked independently from an ACN:$H_2O$ (1:1, v/v) based solution and only up to 5000 ng/L since the expected concentration in wastewater within this range (Equation SI 5).

$$Relative\ recovery\ [\%] = \frac{C_{spiked\ sample} - C_{unspiked\ sample}}{C_{spiked}} * 100 \qquad (SI\ 5)$$

Relative recoveries were only calculated if the concentration in the unspiked sample was lower than twice the spiked concentration. If the concentration in the unspiked sample was lower than twice the LOQ only recoveries from samples with a spiked concentration above four times the LOQ were reported. Recoveries were not considered if the total concentration in the spiked sample exceeded the highest calibration point concentration.

##### SI 1.4.5 Precision

Three aliquots of the same wastewater sample were prepared and analyzed separately in every measurement batch to assess the overall precision of the analytical method.

The precision was computed as the relative standard deviation of the three measurements (Equation SI 6).

$$Relative\ standard\ deviation\ [\%] = \frac{std\ (C_{Replicates})}{mean\ (C_{Replicates})} * 100 \qquad (SI\ 6)$$

#### SI 1.5 Pollen measurement station characteristics

Table SI 4: Characteristics of local pollen measurement station

| Characteristics | Value |
|---|---|
| Station code | PZH |
| Latitude/Longitude | 47.378158 / 8.5658 |
| Station | Zurich |
| Data since | 01.05.1981 |
| Station height | 558 meters above sea level |
| Exposition | South-facing slope |
| Operator | MeteoSwiss |

#### SI 1.6 Pollen load classes

Pollen levels are categorized by MetoSwiss into four load classes: low, moderate, high, and very high, according to the thresholds listed in Table SI 5. This classification was established based on the empirical observations of allergists in the 1990s [1]. Individuals with high sensitivity may experience symptoms even at low pollen levels. As pollen concentration increases, a greater number of individuals are affected, and the severity of symptoms intensifies. Importantly, thresholds vary for different plant species, reflecting the unique allergenic potential of each species' pollen.

Table SI 5: Threshold values for pollen load classes of allergenic pollen types

| Pollen species | low | moderate | high | very high |
|---|---|---|---|---|
| Hazel (Corylus) | 1 - 10 | 11 - 69 | 70 - 249 | ≥ 250 |
| Alder (Alnus) | 1 - 10 | 11 - 69 | 70 - 249 | ≥ 250 |
| Ash (Fraxinus) | 1 - 10 | 11 - 99 | 100 - 349 | ≥ 350 |
| Birch (Betula) | 1 - 10 | 11 - 69 | 70 - 299 | ≥ 300 |
| Hornbeam (Carpinus) | 1 - 10 | 11 - 69 | 70 - 249 | ≥ 250 |
| Plane (Platanus) | 1 - 49 | 50 - 99 | 100 - 399 | ≥ 400 |
| Oak (Quercus) | 1 - 49 | 50 - 129 | 130 - 399 | ≥ 400 |
| Beech (Fagus) | 1 - 49 | 50 - 129 | 130 - 399 | ≥ 400 |
| Chestnut (Castanea) | 1 - 99 | 100 - 199 | 200 - 699 | ≥ 700 |
| Grass (Poaceae) | 1 - 19 | 20 - 49 | 50 - 149 | ≥ 150 |
| Sorrel (Rumex) | 1 - 14 | 15 - 24 | 25 - 59 | ≥ 60 |
| Plantain (Plantago) | 1 - 14 | 15 - 24 | 25 - 59 | ≥ 60 |
| Mugwort (Artemisia) | 1 - 5 | 6 - 14 | 15 - 49 | ≥ 50 |
| Ragweed (Ambrosia) | 1 - 5 | 6 -10 | 11 - 39 | ≥ 40 |

### SI 2. Results and Discussion

#### SI 2.1 Antihistamine substance properties

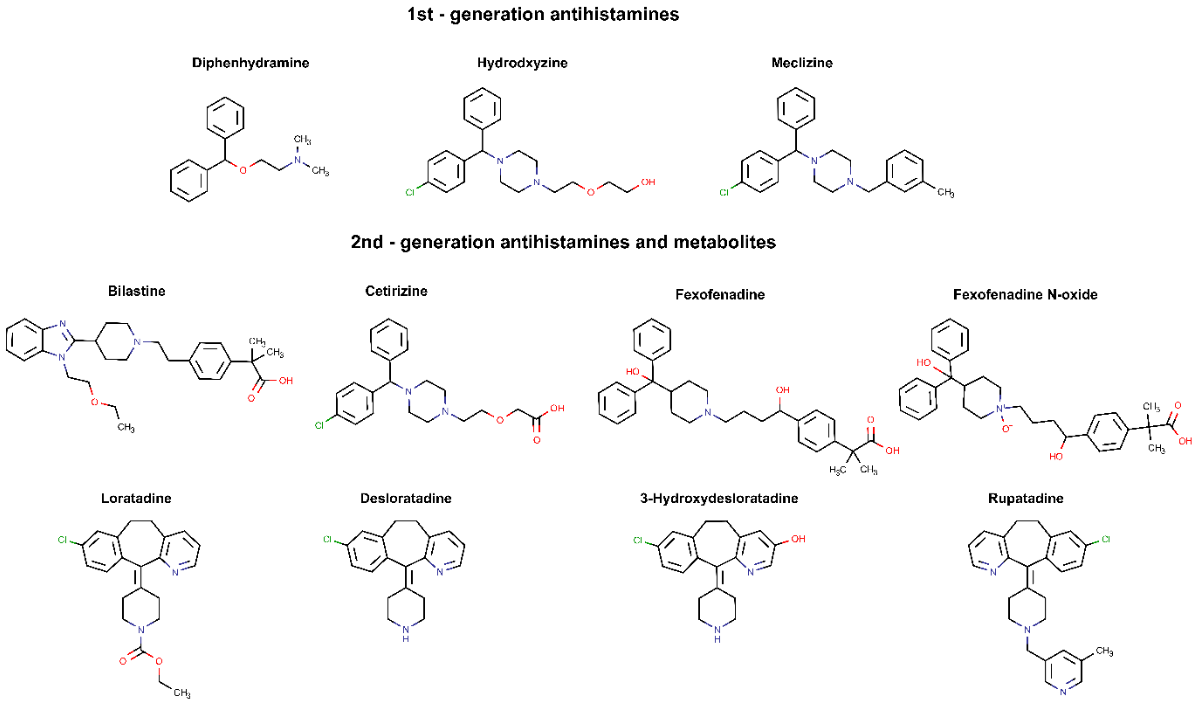


Figure SI 2: Molecular structures of antihistamine markers (created with Marvin, Chemaxon)

Table SI 6: Pharmacological properties of analyzed substances; maximum daily dose, excretion, and plasma half-life.

| Substance | Antihistamine type | Daily Dose [mg][2] | Unchanged Urinary Excretion [%] | Plasma elimination time $t_{1/2}$ [h] | Remark |
|---|---|---|---|---|---|
| 3-Hydroxydesloratadine (3-OH-DL) | metabolite | - | - | - | - |
| Bilastine | 2nd generation | 10 | 33 [3] | 14.5 [4] | 67% excreted unchanged in feces [3] |
| Cetirizine | 2nd generation | 10 (*2HCl) | 60 [5] | 8 [6] | Levocetirizine eutomer of cetrizine. 10% of dosis excreted in feces (primarily unchanged) [5] |
| Desloratadine | 2nd generation | 5 | 1.7 [7] | 24 [8] | urinary excretion: 22% as 3-OH-DL-glucuronide, 0.4% as 3-OH-D, fecal excretion: 28.4% as 3-hydroxydesloratadine, 6.7% as desloratadine [7] |
| Diphenhydramine | 1st generation | 50 (*HCl) | 3 [9] | 14 [9] | - |
| Fexofenadine | 2nd generation | 180 (*HCl) | 11 [10] | 11-15 [6] | Only little metabolization. 80% of radioactive dose excreted in feces [6] |
| Fexofenadine N-oxide | impurity | - | - | - | - |
| Hydroxyzine | 1st generation | 100 (*HCl) | NA | NA | 70% excreted as cetirizine in urine [11] |
| Loratadine | 2nd generation | 10 | <1 [12] | 16h [12] | Urinary excretion: 2% as desloratadine, 17% as 3-hydroxy-desloratadine glucuonide. Fecal excretion: 17% as 3-hydroxy-desloratadine, 5.4% as loratadine, 2.7% as desloratadine [12] |
| Meclizine | 1st generation | 25-50 (*2HCl) | NA | 5-6 [13] | Excreted in urine as metabolite and in feces as unchanged drug [13] |
| Rupatadine | 2nd generation | no registered products in CH | NA | 7 [14] | Metabolization to desloratadine and 3-hydroxydesloratadine |

#### SI 2.2 LCMS Quantification

Table SI 7: Summary of LCMS Analytics Quality Control and Quantification. The data is presented for individual measurement batches, with quality control parameters determined as described above. (m/z: Mass to charge ratio, RT: Retention time, LOQ: Limit of quantification, SD: Standard deviation, NA: Not available)

| Substance | Measurement batch ID | m/z theoretical [M+H]+ | m/z delta [ppm] | RT measured [min] | SD RT [min] | LOQ [ng/L] | Sample conc. [ng/L] | Samples above LOQ [%] | Absolute recovery | Relative recovery [%] | Precision [%] | Number of samples |
|---|---|---|---|---|---|---|---|---|---|---|---|---|
| 3-Hydroxydesloratadine | 220803 | 327.1259 | 0.1609 | 13.4 | 0.13 | 100 | NA | 0 | 0.73 | 118.6 | NA | 78 |
| 3-Hydroxydesloratadine | 221021 | 327.1259 | -0.0227 | 13.1 | 0.03 | 100 | NA | 0 | 0.79 | 109.7 | NA | 182 |
| 3-Hydroxydesloratadine | 230109 | 327.1259 | 0.6757 | 12.9 | 0.06 | 100 | NA | 0 | 0.64 | 100 | NA | 7 |
| Bilastine | 220803 | 464.2908 | -0.2132 | 14.1 | 0.06 | 25 | 135 | 94.9 | 0.83 | 96.4 | 1.5 | 78 |
| Bilastine | 221021 | 464.2908 | -0.2179 | 13.8 | 0.02 | 45 | 137 | 81.9 | 0.86 | 96.7 | 3.8 | 182 |
| Bilastine | 230109 | 464.2908 | -0.0265 | 13.7 | 0.03 | 50 | 80 | 100 | 0.85 | 100.3 | NA | 7 |
| Bilastine | 230424 | 464.2908 | 0.2348 | 13.8 | 0 | 25 | 155 | 100 | 0.86 | 98.4 | NA | 7 |
| Bilastine | 230711 | 464.2908 | -0.3419 | 14.7 | 0.02 | 50 | 399 | 100 | 0.8 | 103.3 | NA | 6 |
| Bilastine | 231106 | 464.2908 | 0.9121 | 13.6 | 0.03 | 25 | 138 | 100 | 0.88 | 89.3 | 22.8 | 8 |
| Bilastine | 240131 | 464.2908 | -0.3505 | 13.9 | 0.12 | 25 | 93 | 85.7 | 1 | 106.9 | 14.6 | 7 |
| Cetirizine | 220803 | 389.1626 | -0.0469 | 17.2 | 0.02 | 25 | 120 | 91 | 0.73 | 104.3 | 3.8 | 78 |
| Cetirizine | 221021 | 389.1627 | 0.1394 | 17 | 0.04 | 25 | 99 | 90.7 | 0.86 | 101.5 | 8.8 | 182 |
| Cetirizine | 230109 | 389.1626 | 0.5165 | 16.9 | 0.03 | 30 | 58 | 100 | 0.94 | 98.9 | NA | 7 |
| Cetirizine | 230424 | 389.1627 | 0.3105 | 17 | 0 | 10 | 90 | 100 | 0.75 | 98.8 | 3.5 | 7 |
| Cetirizine | 230711 | 389.1626 | 0.3886 | 17.3 | 0 | 10 | 227 | 100 | 0.54 | 100.5 | 3.1 | 6 |
| Cetirizine | 231106 | 389.1626 | 1.4596 | 17 | 0 | 10 | 84 | 100 | 0.69 | 99.7 | 1.4 | 8 |
| Cetirizine | 240131 | 389.1626 | -0.1612 | 17.3 | 0 | 10 | 58 | 85.7 | 0.72 | 99 | 3.6 | 7 |
| Desloratadine | 220803 | 311.1309 | 0.026 | 13.7 | 0.11 | 100 | NA | 0 | 0.61 | 107.4 | NA | 78 |
| Desloratadine | 221021 | 311.1309 | 0.1195 | 13.4 | 0.05 | 500 | NA | 0 | 0.7 | 101.9 | NA | 182 |
| Desloratadine | 230109 | 311.1309 | 0.7553 | 13.4 | 0.11 | 500 | NA | 0 | 0.65 | 101.2 | NA | 7 |
| Diphenhydramine | 220803 | 256.1696 | -0.1127 | 14.4 | 0.1 | 10 | 48 | 100 | 0.77 | 94.4 | 7.1 | 78 |
| Diphenhydramine | 221021 | 256.1696 | 0.2016 | 14.1 | 0.06 | 25 | 53 | 78 | 0.8 | 97.7 | 7.4 | 182 |
| Diphenhydramine | 230109 | 256.1696 | 0.4183 | 13.9 | 0.07 | 25 | 49 | 100 | 0.89 | 99.4 | NA | 7 |
| Diphenhydramine | 230424 | 256.1696 | 0.1151 | 14 | 0 | 25 | 54 | 100 | 0.87 | 102.3 | NA | 7 |
| Diphenhydramine | 230711 | 256.1696 | -0.0955 | 14.5 | 0.08 | 10 | 61 | 100 | 0.84 | 99.5 | NA | 6 |
| Diphenhydramine | 231106 | 256.1696 | 0.8386 | 13.9 | 0.04 | 25 | 62 | 100 | 0.9 | 92.2 | 2.2 | 8 |
| Diphenhydramine | 240131 | 256.1696 | 0.7334 | 14.1 | 0.15 | 25 | 79 | 28.6 | 0.88 | 97.7 | 13.8 | 7 |
| Fexofenadine | 220803 | 502.2952 | -0.0455 | 15.5 | 0.12 | 25 | 1392 | 100 | 0.9 | 104.7 | 3.7 | 78 |
| Fexofenadine | 221021 | 502.2952 | -0.1054 | 15.2 | 0.06 | 25 | 1312 | 100 | 0.96 | 97.4 | 3.2 | 182 |
| Fexofenadine | 230109 | 502.2952 | 0.1303 | 15 | 0.08 | 100 | 446 | 100 | 0.98 | 91.4 | 10.2 | 7 |
| Fexofenadine | 230424 | 502.2952 | 0.1001 | 15 | 0 | 50 | 1060 | 100 | 0.91 | 96.6 | 2.8 | 7 |
| Fexofenadine | 230711 | 502.2952 | -0.6874 | 15.8 | 0.13 | 50 | 4354 | 100 | 0.88 | 100.3 | 3 | 6 |
| Fexofenadine | 231106 | 502.2952 | 0.8546 | 15.1 | 0.04 | 50 | 997 | 100 | 0.94 | 91.5 | 1.3 | 8 |
| Fexofenadine | 240131 | 502.2952 | -0.3363 | 15.2 | 0.14 | 50 | 464 | 100 | 1.16 | 103.6 | 2.1 | 7 |
| Fexofenadine N-oxide | 220803 | 518.2901 | -1.8902 | 15.8 | 0.19 | 100 | NA | 0 | 0.84 | 80.8 | NA | 78 |
| Fexofenadine N-oxide | 221021 | 518.2901 | -0.288 | 15.3 | 0.04 | 100 | NA | 0 | 0.84 | 91 | NA | 182 |
| Fexofenadine N-oxide | 230109 | 518.2901 | -0.3467 | 15.2 | 0.12 | 100 | NA | 0 | 0.74 | 94.6 | NA | 7 |
| Hydroxyzine | 220803 | 375.1834 | -0.053 | 15.7 | 0.17 | 25 | NA | 0 | 0.75 | 100 | NA | 78 |
| Hydroxyzine | 221021 | 375.1834 | -0.0912 | 15.4 | 0.05 | 25 | NA | 0 | 0.89 | 98.8 | NA | 182 |
| Hydroxyzine | 230109 | 375.1834 | 0.2415 | 15.3 | 0.13 | 25 | NA | 0 | 0.88 | 97.3 | NA | 7 |
| Loratadine | 220803 | 383.1521 | -0.8398 | 19.1 | 0.18 | 50 | NA | 0 | 0.89 | 110.4 | NA | 78 |
| Loratadine | 221021 | 383.1521 | -0.1612 | 19.1 | 0.11 | 50 | NA | 0 | 0.83 | 101.9 | NA | 182 |
| Loratadine | 230109 | 383.1521 | -0.3573 | 18.8 | 0.18 | 50 | NA | 0 | 0.74 | 101.5 | NA | 7 |

| Substance | Measurement batch ID | m/z theoretical [M+H]+ | m/z delta [ppm] | RT measured [min] | SD RT [min] | LOQ [ng/L] | Sample conc. [ng/L] | Samples above LOQ [%] | Absolute recovery | Relative recovery [%] | Precision [%] | Number of samples |
|---|---|---|---|---|---|---|---|---|---|---|---|---|
| Meclozin | 220803 | 391.1936 | 0.1026 | 17.2 | 0.16 | 100 | NA | 0 | 0.56 | 118.7 | NA | 78 |
| Meclozin | 221021 | 391.1936 | 0.0744 | 16.8 | 0.07 | 250 | NA | 0 | 0.57 | 105.2 | NA | 182 |
| Meclozin | 230109 | 391.1936 | 0.751 | 16.8 | 0.13 | 100 | NA | 0 | 0.74 | 96 | NA | 7 |
| Rupatadine | 220803 | 416.1888 | 0.3606 | 14.7 | 0.19 | 1000 | NA | 0 | 0.78 | 102.3 | NA | 78 |
| Rupatadine | 221021 | 416.1888 | 0.1191 | 14.3 | 0.06 | 1000 | NA | 0 | 0.79 | 99.2 | NA | 182 |
| Rupatadine | 230109 | 416.1888 | -0.3362 | 14.2 | 0.12 | 1000 | NA | 0 | 0.89 | 89.1 | NA | 7 |

#### SI 2.3 Airborne pollen patterns and fexofenadine wastewater patterns

Additional temporal patterns of pollen classified as allergenic or for which there are indications of allergenicity are displayed below.

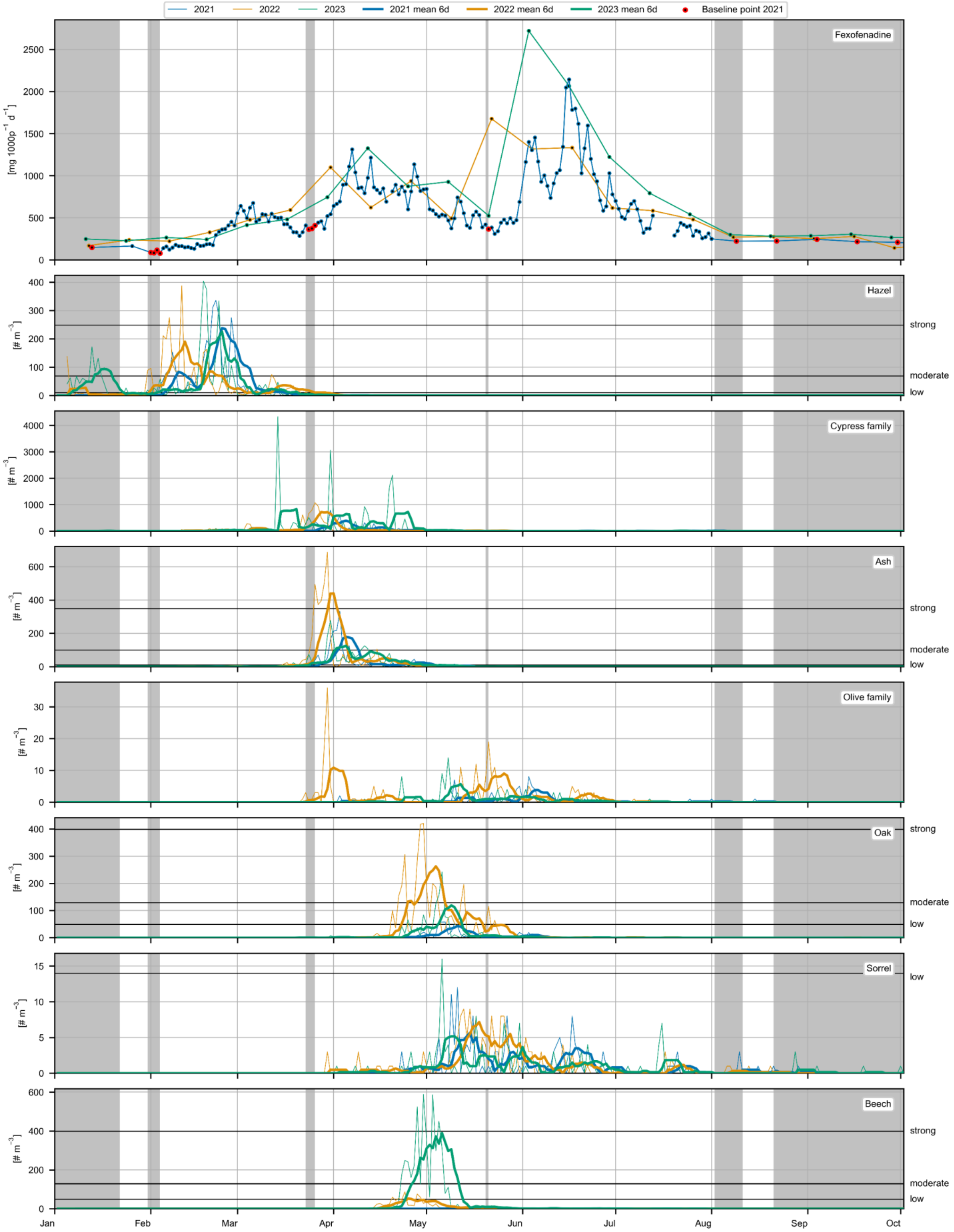


Figure SI 3: Concentrations of allergenic airborne pollen (number of pollen grains/m³) and fexofenadine wastewater loads (mg/day/1000 people) in Zurich during 2021, 2022, and 2023. Upper limits of pollen taxa-specific load classes are indicated by horizontal lines. Grey shading indicates days in 2021 when no pollen taxa exceeded the lowest load class in the previous nine days, with red circles highlighting the corresponding fexofenadine loads as baseline points. Thin lines show actual observed pollen concentrations, while thick lines represent the mean pollen values of the previous six days used for modeling.

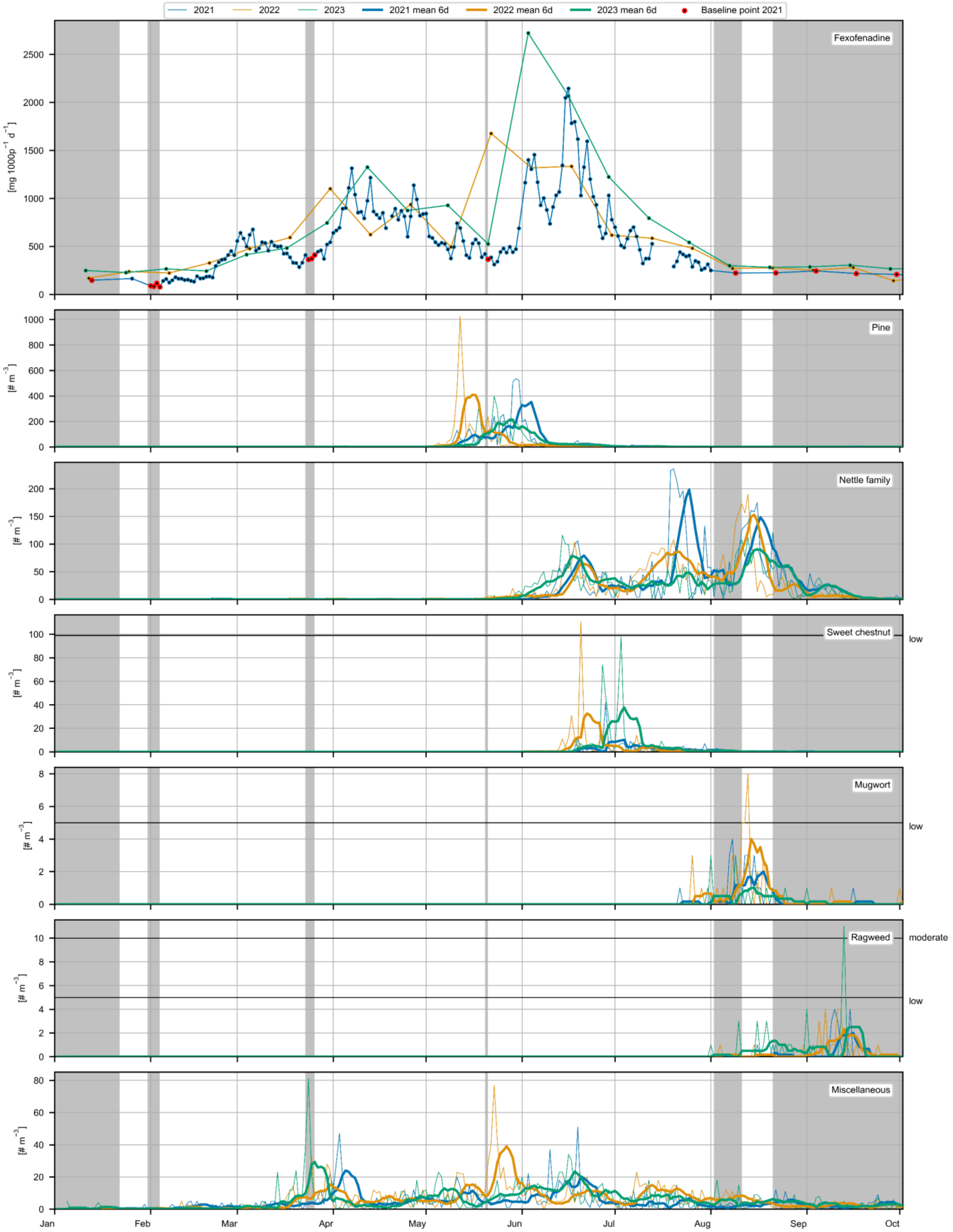


Figure SI 4: Concentrations of allergenic airborne pollen (number of pollen grains/m³) and fexofenadine wastewater loads (mg/day/1000 people) in Zurich during 2021, 2022, and 2023. Upper limits of pollen taxa-specific load classes are indicated by horizontal lines. Grey shading indicates days in 2021 when no pollen taxa exceeded the lowest load class in the previous nine days, with red circles highlighting the corresponding fexofenadine loads as baseline points. Thin lines show actual observed pollen concentrations, while thick lines represent the mean pollen values of the previous six days used for modeling.

Additional temporal patterns of pollen classified as non- allergenic are displayed below.

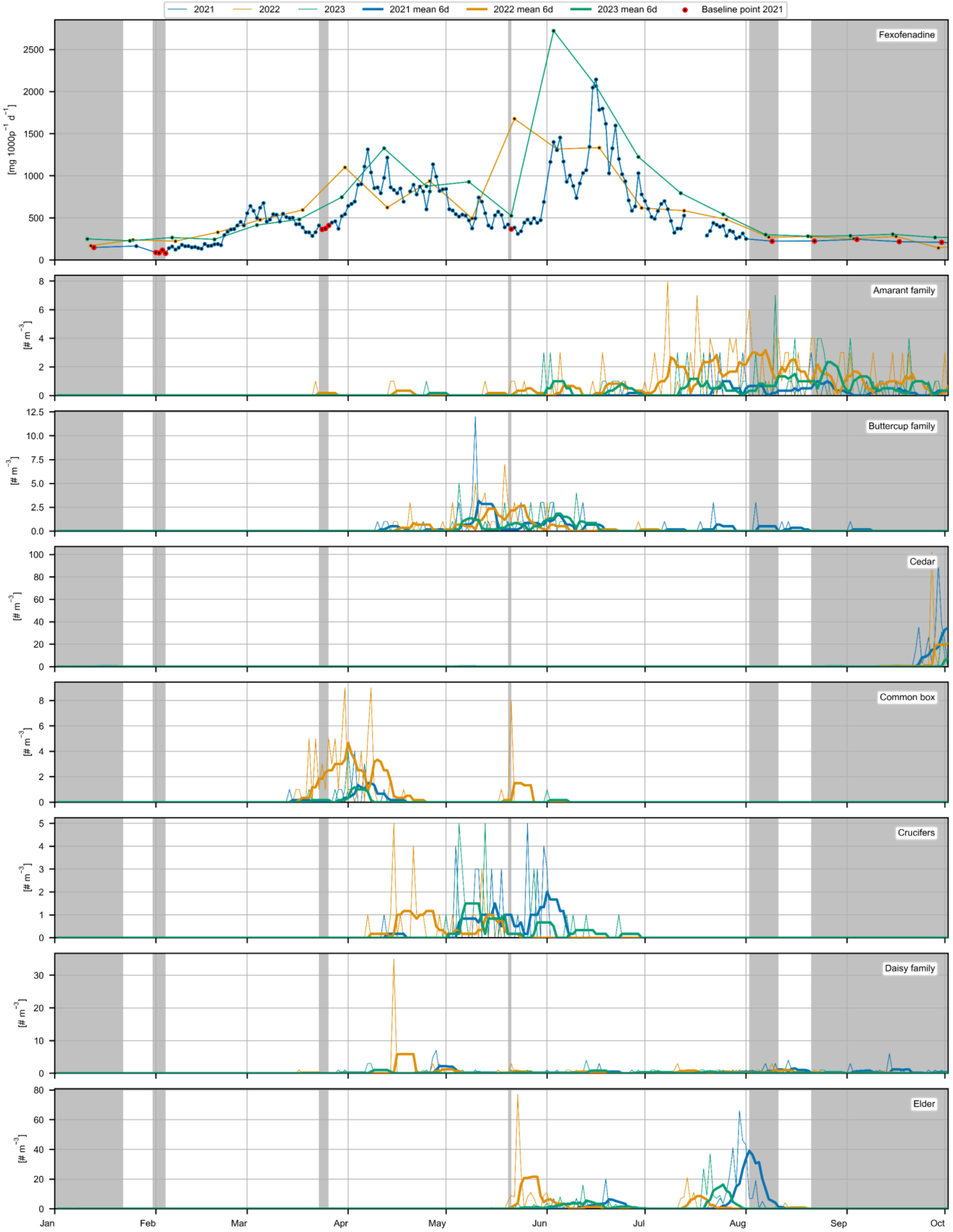


Figure SI 5: Concentrations of allergenic airborne pollen (number of pollen grains/m³) and fexofenadine wastewater loads (mg/day/1000 people) in Zurich during 2021, 2022, and 2023. Upper limits of pollen taxa-specific load classes are indicated by horizontal lines. Grey shading indicates days in 2021 when no pollen taxa exceeded the lowest load class in the previous nine days, with red circles highlighting the corresponding fexofenadine loads as baseline points. Thin lines show actual observed pollen concentrations, while thick lines represent the mean pollen values of the previous six days used for modeling.

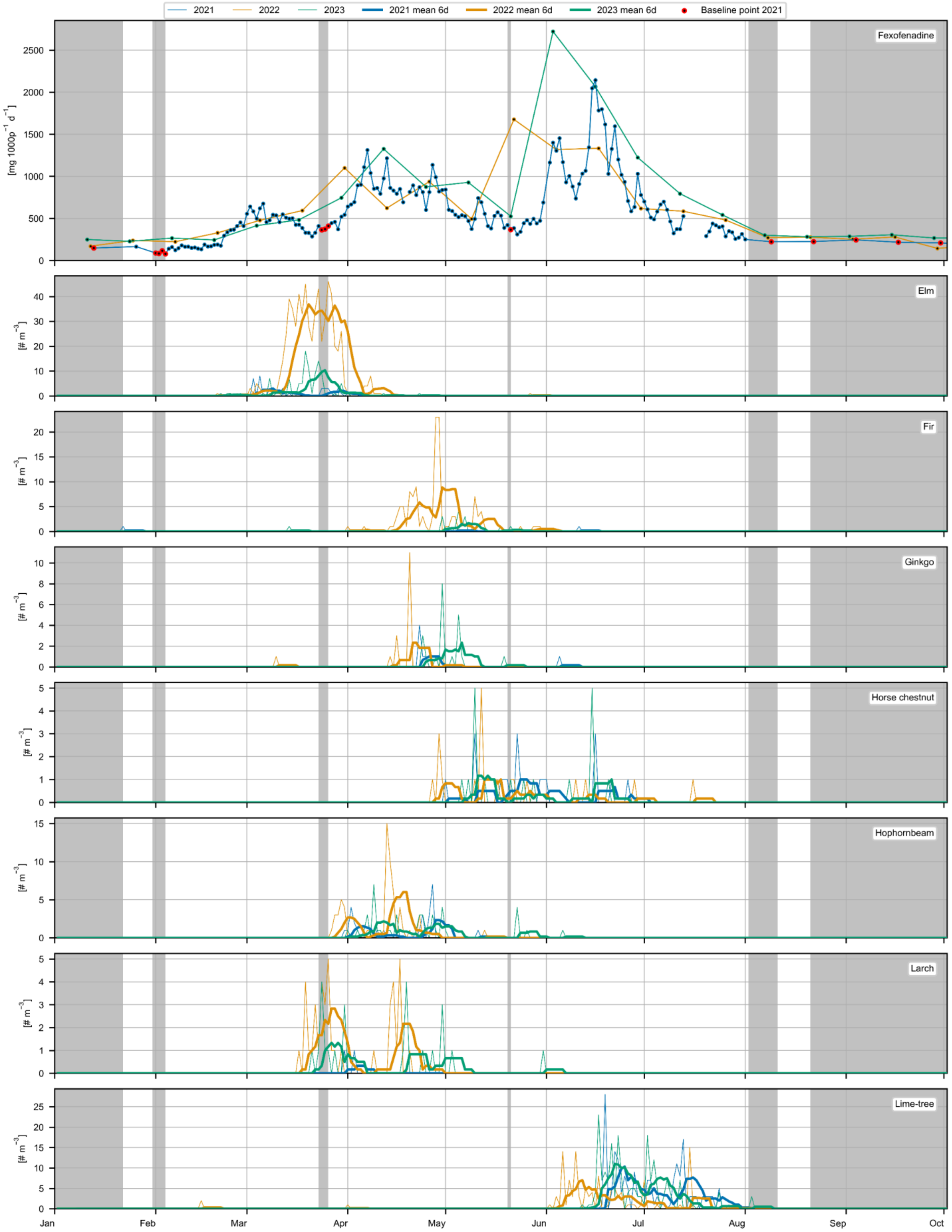


Figure SI 6: Concentrations of allergenic airborne pollen (number of pollen grains/m³) and fexofenadine wastewater loads (mg/day/1000 people) in Zurich during 2021, 2022, and 2023. Upper limits of pollen taxa-specific load classes are indicated by horizontal lines. Grey shading indicates days in 2021 when no pollen taxa exceeded the lowest load class in the previous nine days, with red circles highlighting the corresponding fexofenadine loads as baseline points. Thin lines show actual observed pollen concentrations, while thick lines represent the mean pollen values of the previous six days used for modeling.

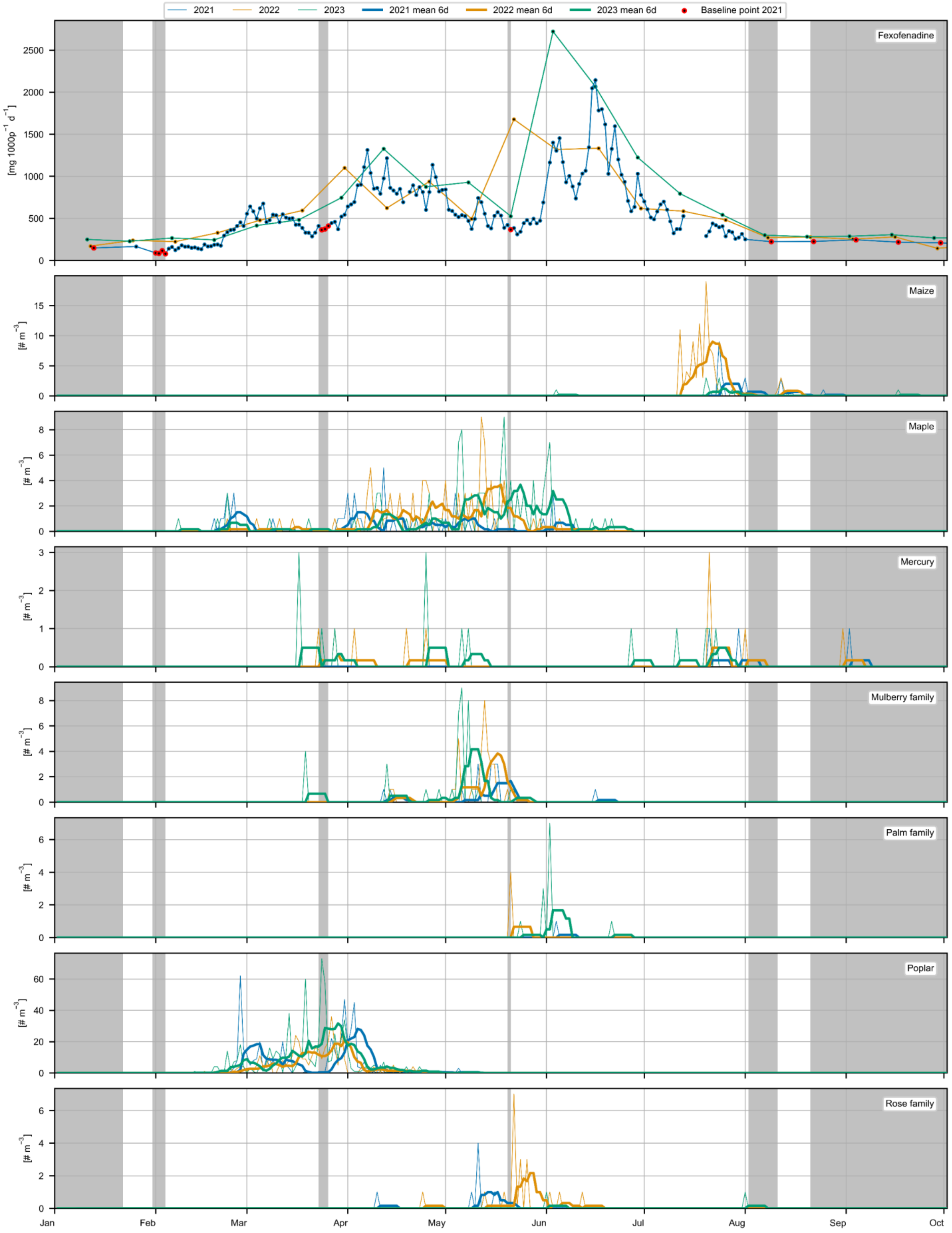


Figure SI 7: Concentrations of allergenic airborne pollen (number of pollen grains/m³) and fexofenadine wastewater loads (mg/day/1000 people) in Zurich during 2021, 2022, and 2023. Upper limits of pollen taxa-specific load classes are indicated by horizontal lines. Grey shading indicates days in 2021 when no pollen taxa exceeded the lowest load class in the previous nine days, with red circles highlighting the corresponding fexofenadine loads as baseline points. Thin lines show actual observed pollen concentrations, while thick lines represent the mean pollen values of the previous six days used for modeling.

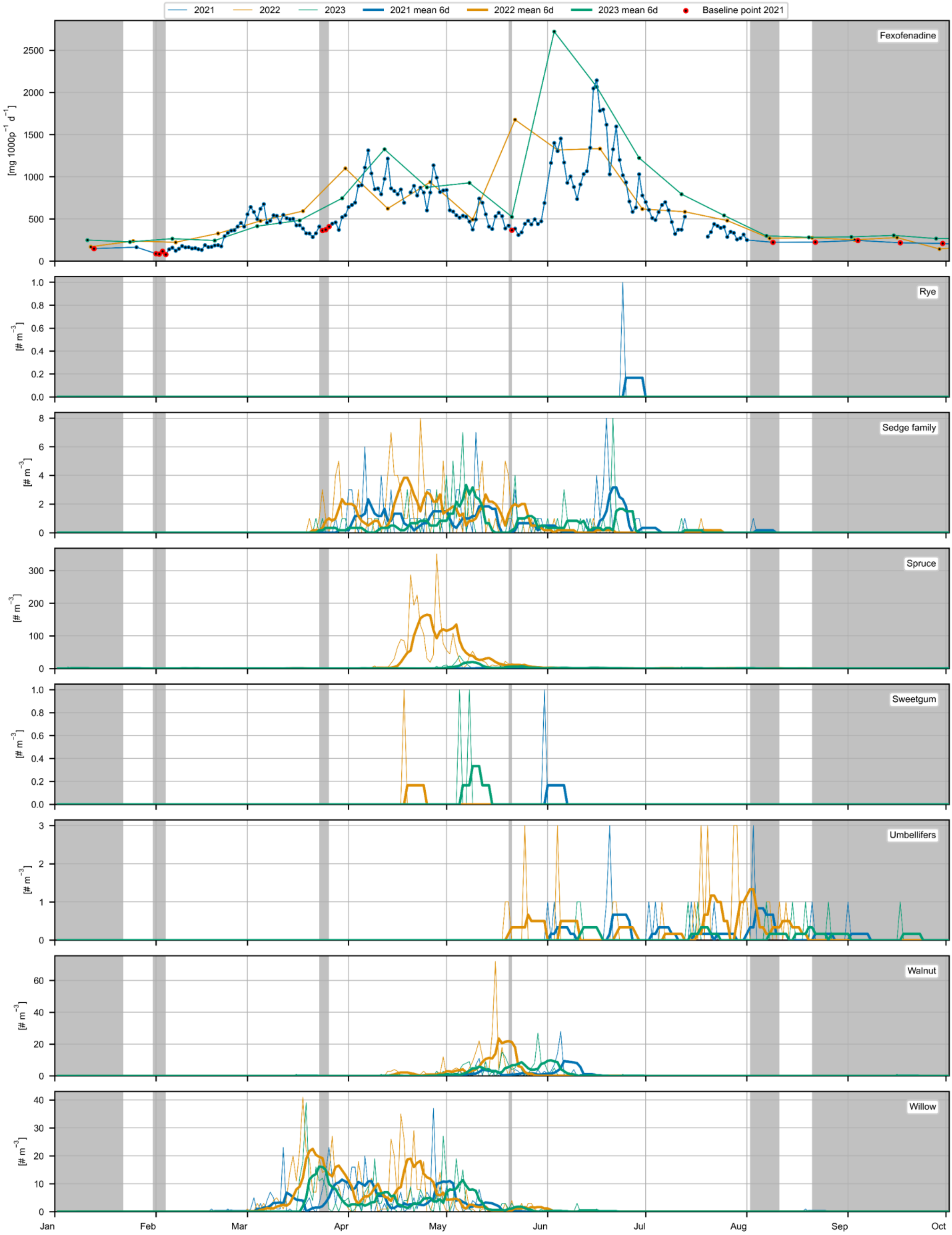


Figure SI 8: Concentrations of allergenic airborne pollen (number of pollen grains/m³) and fexofenadine wastewater loads (mg/day/1000 people) in Zurich during 2021, 2022, and 2023. Upper limits of pollen taxa-specific load classes are indicated by horizontal lines. Grey shading indicates days in 2021 when no pollen taxa exceeded the lowest load class in the previous nine days, with red circles highlighting the corresponding fexofenadine loads as baseline points. Thin lines show actual observed pollen concentrations, while thick lines represent the mean pollen values of the previous six days used for modeling.

#### SI 2.4 Pollen antihistamine model

##### SI 2.4.1 Pollen concentration transformation

Our data indicates a delayed and smoothened relation between airborne pollen concentrations and wastewater fexofenadine loads. Different transformations were applied to the pollen concentrations to understand the correlation of pollen exposure and antihistamine consumption at population scale.

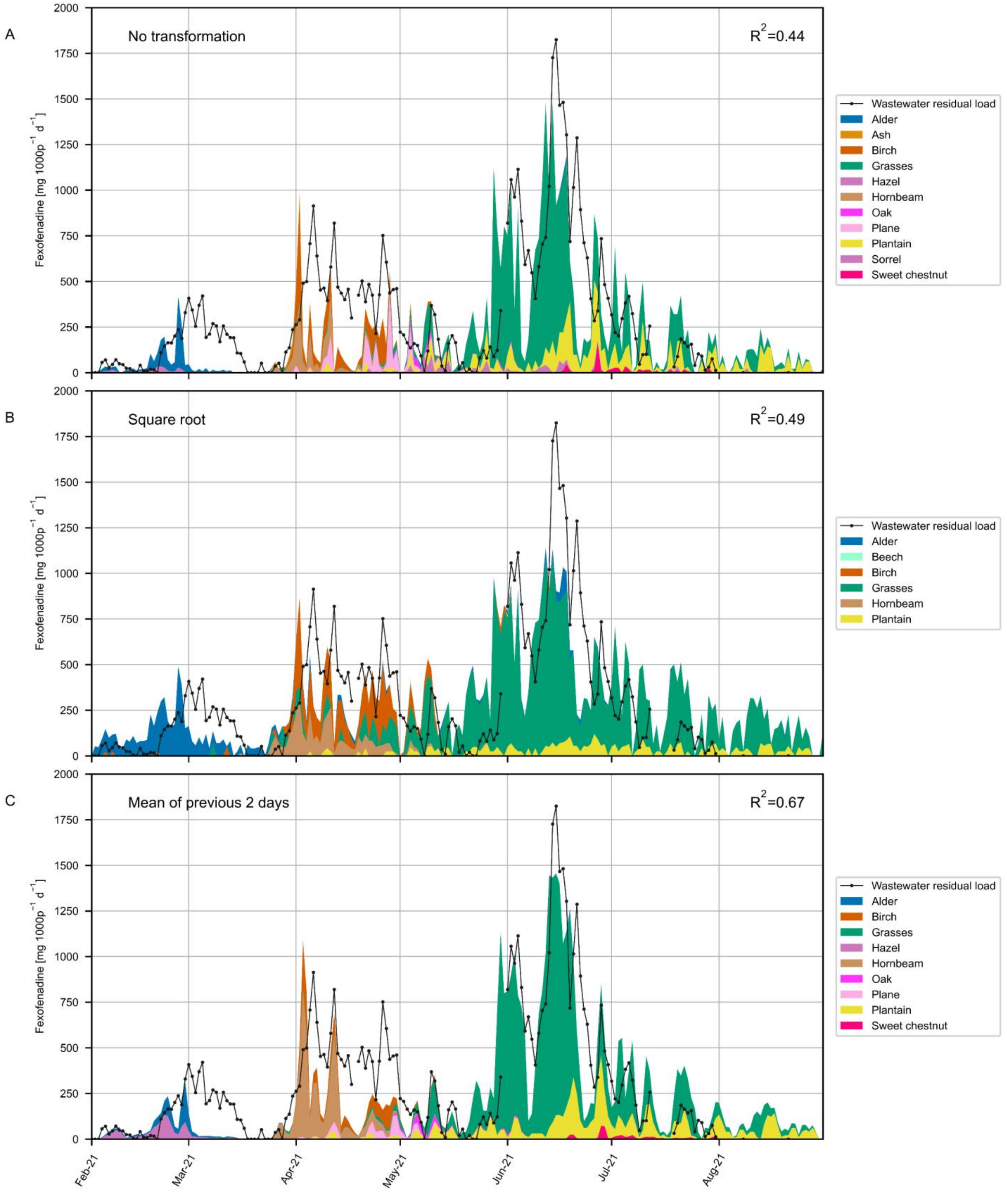


Figure SI 9: Influence of pollen concentration transformations on NNLS modeling of fexofenadine consumption. The relationship between transformed pollen concentrations and observed fexofenadine consumption was assessed based on residual fexofenadine loads (baseline consumption excluded) from 2021 to 2023, along with the pollen concentration of the 14 allergenic pollen taxa. Only values from 2021 are shown for better visualization. In (A), the pollen concentrations remained unmodified. In (B), the square root transformation was applied to the pollen concentration, and in (C), a two-day moving average of preceding days was used. The coefficient of determination (R2) was calculated to determine model performance.

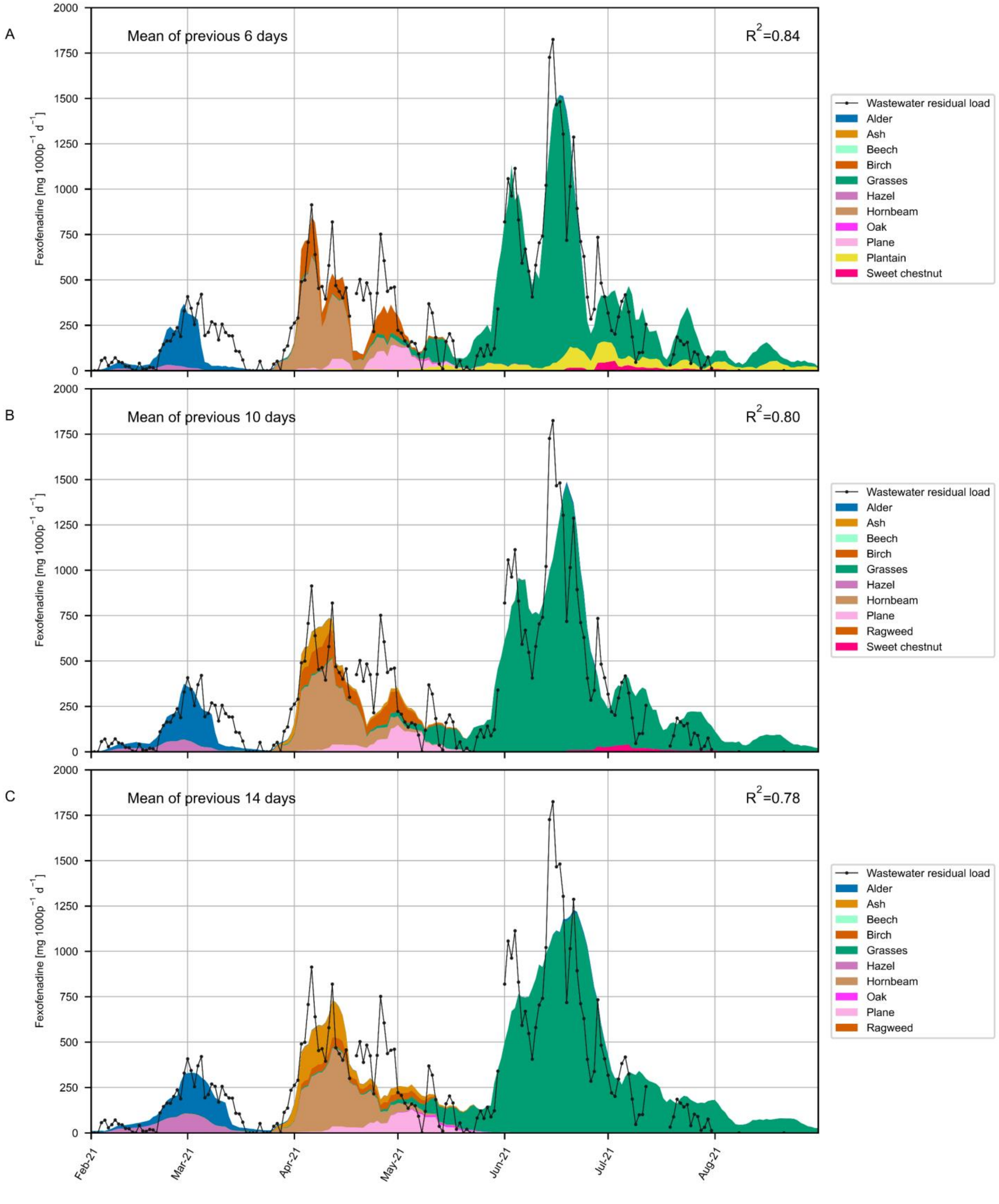


Figure SI 10: Influence of pollen concentration transformations on NNLS modeling of fexofenadine consumption. The relationship between transformed pollen concentrations and observed fexofenadine consumption was assessed based on residual fexofenadine loads (baseline consumption excluded) from 2021 to 2023, along with the pollen concentration of the 14 allergenic pollen taxa. Only values from 2021 are shown for better visualization. In (A), a six-day moving average of preceding days was used. In (B), the average pollen concentration of the previous 10 days, and in (C), the average pollen concentration of the previous 14 days was employed to determine the relationship.

Among the investigated pollen concentration modifications, the consumption of fexofenadine was best described by the average pollen concentrations of the previous six days ($R^2$=0.84).

##### SI 2.4.2 Linearity of pollen antihistamine correlation

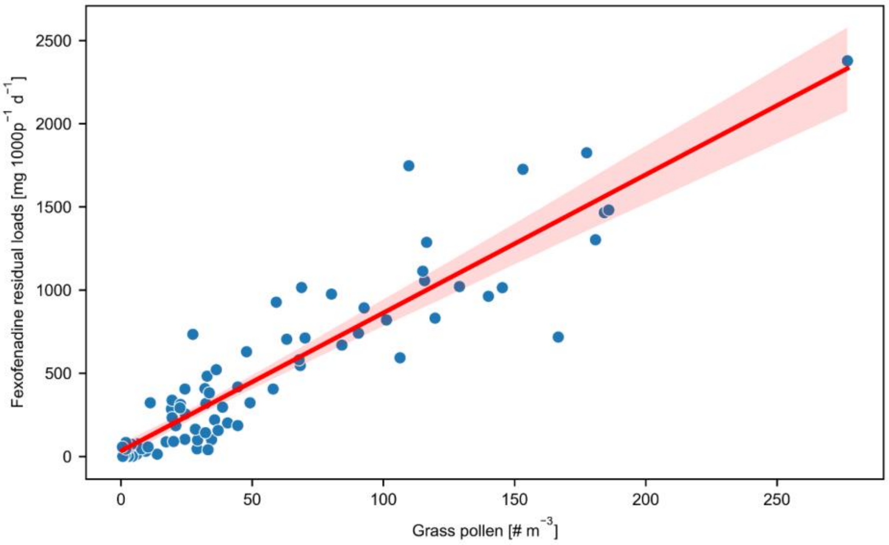


Figure SI 11: Scatter plot with regression line showing the relationship between grass pollen concentrations and baseline-subtracted fexofenadine residual loads from June to September of 2021 to 2023. During this period, grass is the dominant allergenic pollen present, and interference from other pollen taxa is minimal, making it suitable to demonstrate the linear relationship between observed fexofenadine residual loads and grass pollen concentrations.

##### SI 2.4.3 Multivariate linear least squares analysis.

The multicollinearity among pollen taxa poses significant challenges in modeling. To address this, we employed different sets of pollen taxa as explanatory variables in both Non-Negative Least Squares (NNLS) and Ordinary Least Squares (OLS) regression models. Despite the strong overall fit indicated by the high R-squared value, diagnostic plots revealed signs of heteroscedasticity and non-normality in the residuals. These issues are primarily attributed to the nature of the data, particularly the overrepresentation of low pollen concentration events, which can lead to overfitting. Additionally, during different pollen periods, especially the grass pollen period, the model captures the general trend but exhibits large errors between predicted and observed values (e.g. 19.6.2024, 15.6.2023) due to the significantly higher pollen burdens compared to other periods like the tree pollen season. This discrepancy introduces non-normality in the model residuals, as evidenced by the diagnostic plots. While the model requires improvements to better describe these relationships, the primary aim of this pilot study is to demonstrate that second antihistamine loads correlate well with airborne pollen concentrations.

Further refinement in modeling should be pursued in future studies with more daily measurements. Based on the diagnostic plots and statistical tests, we consider the model sufficient for our descriptive analysis, with p-values interpreted as references. The identified issues do not invalidate the overall trends and relationships captured by the model, but they suggest that caution is warranted when interpreting specific coefficients, particularly those with marginal significance.

#### Non-negative least squares (NNLS) analyses diagnostics

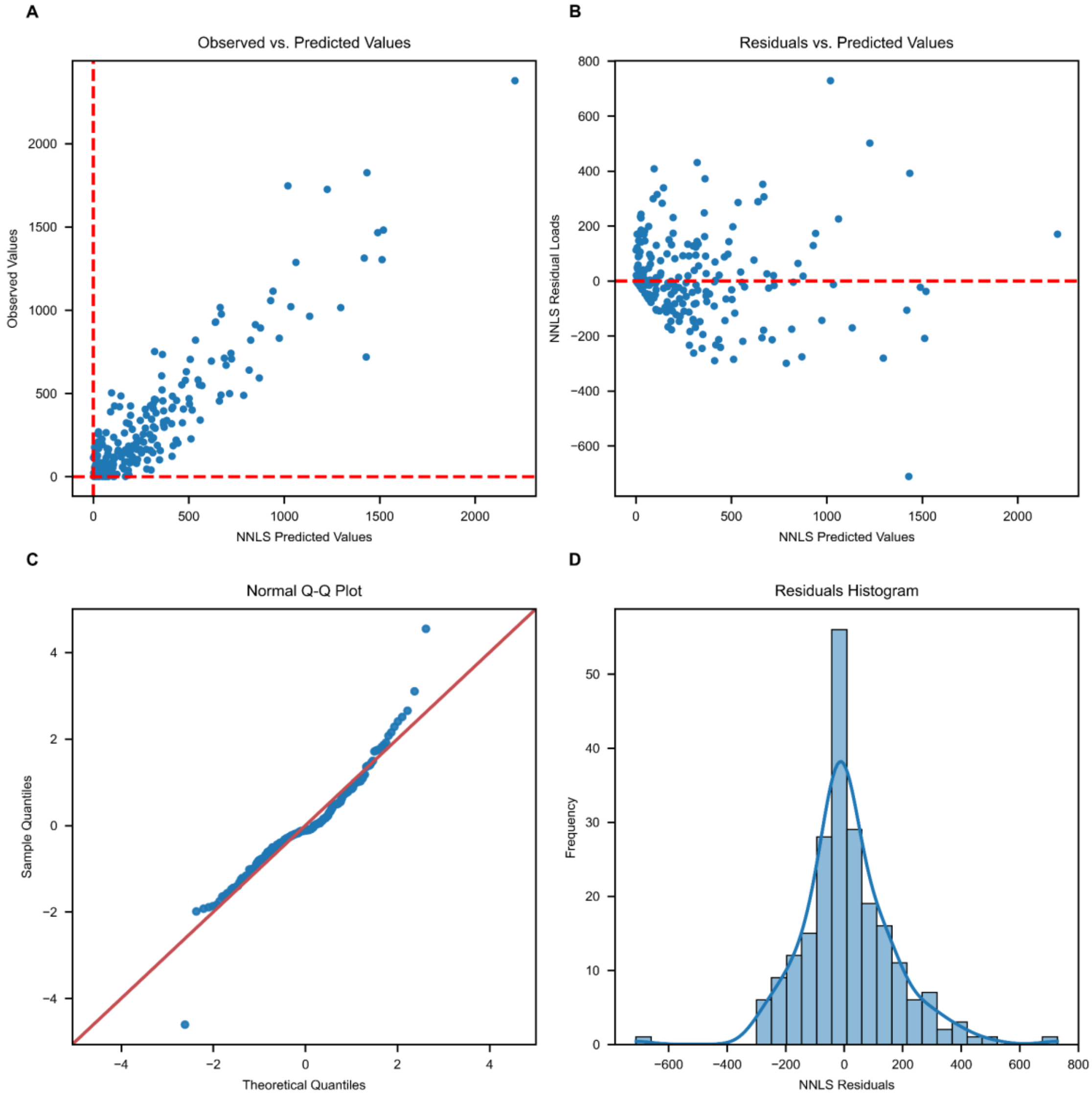


Figure SI 12: Diagnostic plots for the Non-Negative Least Squares (NNLS) regression analysis using airborne pollen concentrations of 14 allergenic taxa as predictors and baseline-subtracted fexofenadine loads as the response variable. The figure includes: (A) Observed vs. Predicted Values, showing the relationship between observed and predicted fexofenadine loads; (B) Residuals vs. Predicted Values, displaying the residuals against predicted values; (C) Normal Q-Q Plot, assessing the normality of residuals; and (D) Residuals Histogram, illustrating the distribution of residuals with a fitted kernel density estimate.

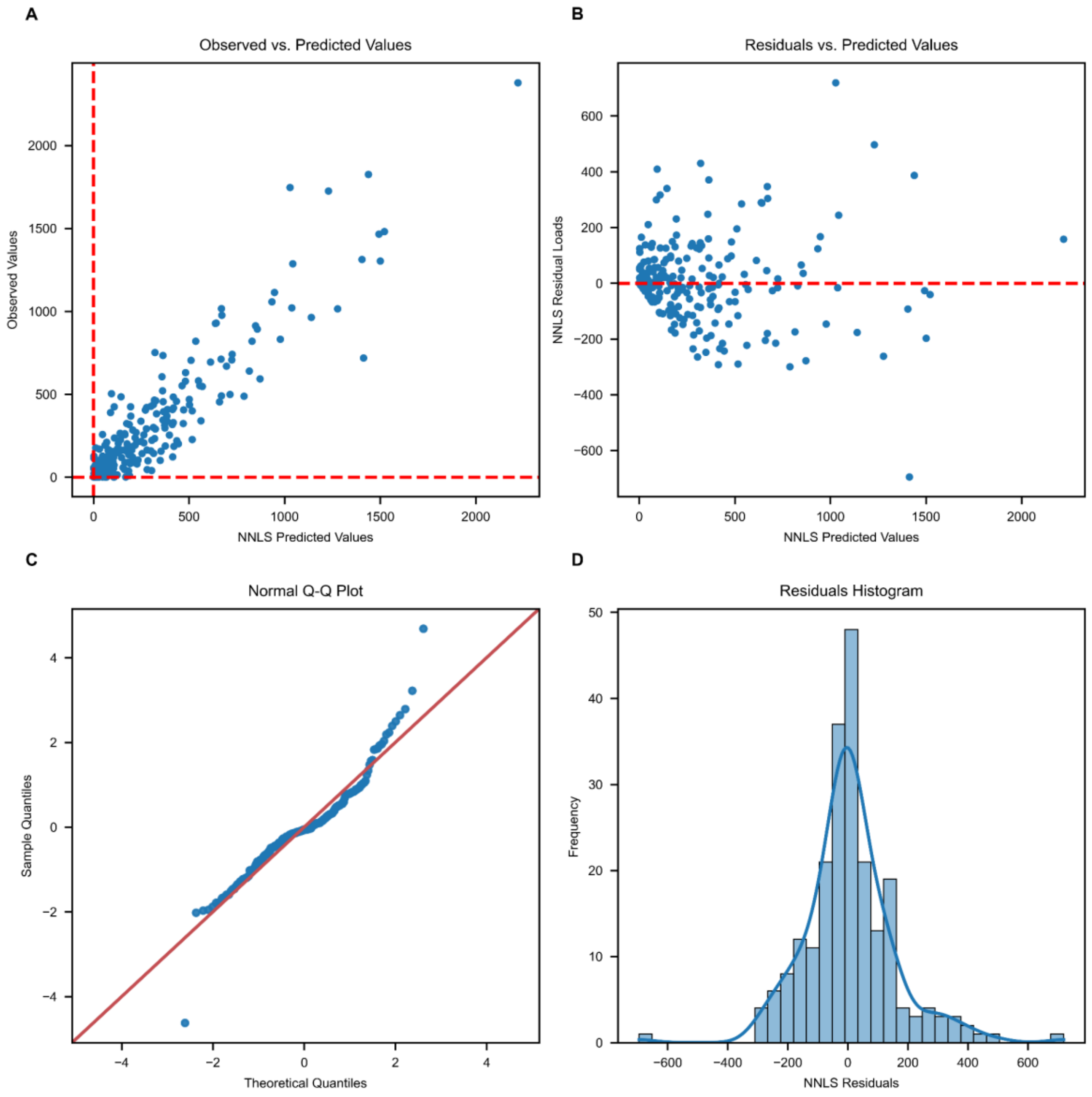


Figure SI 13: Diagnostic plots for the Non-Negative Least Squares (NNLS) regression analysis using airborne pollen concentrations of 14 allergenic taxa as predictors and baseline-subtracted fexofenadine loads as the response variable. The figure includes: (A) Observed vs. Predicted Values, show ing the relationship betw een observed and predicted fexofenadine loads; (B) Residuals vs. Predicted Values, displaying th e residuals against predicted values; (C) Normal Q-Q Plot, assessing the normality of residuals; and (D) Residuals Histogram, illustrating the distribution of residuals w ith a fitted kernel density estimate.

#### Ordinary Least Squares (OLS) analyses

##### OLS with 14 allergenic pollen taxa

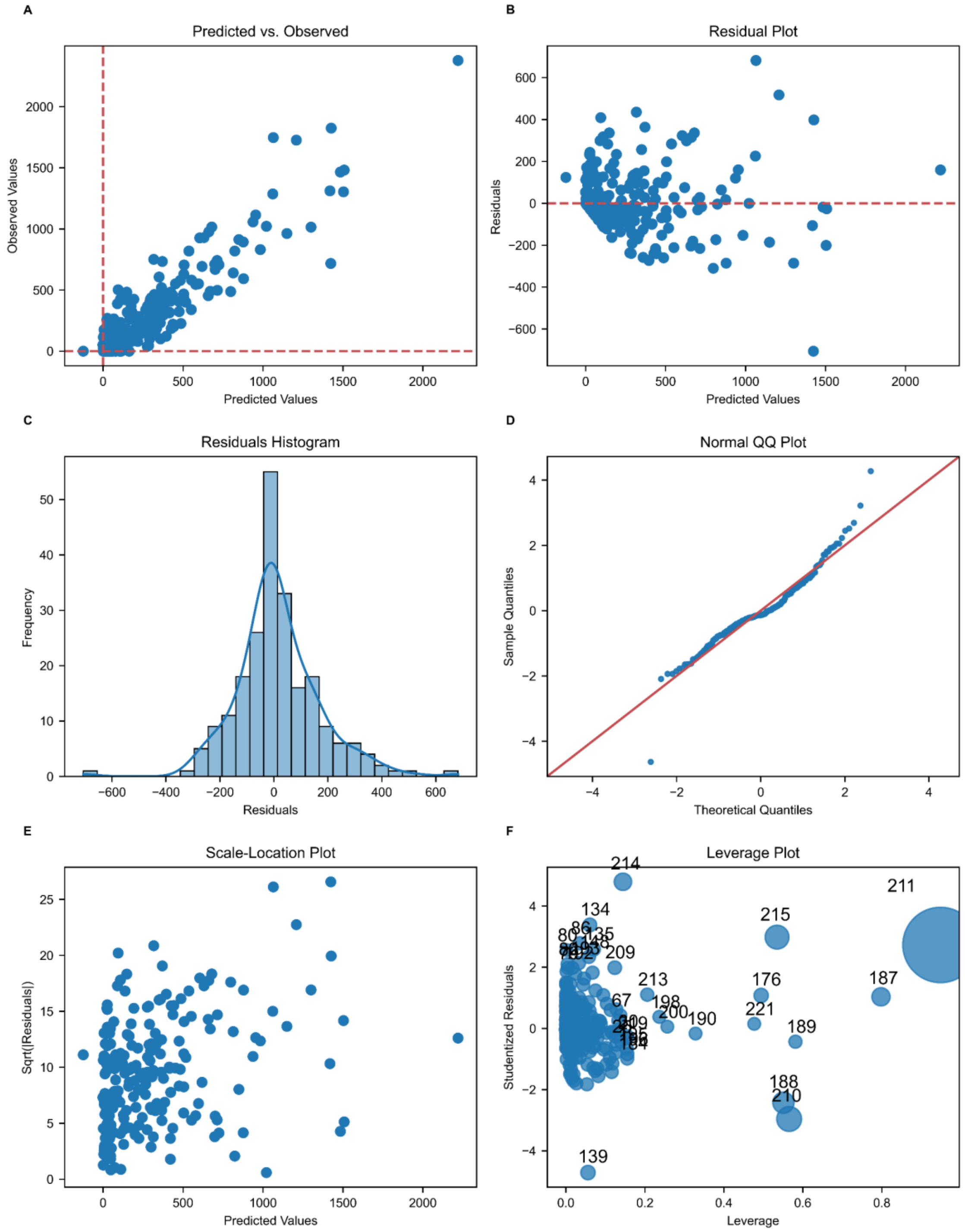


Figure SI 14: Diagnostic plots for the Ordinary Least Squares (OLS) regression analysis using airborne pollen concentrations of 14 allergenic taxa as predictors and baseline-subtracted fexofenadine loads as the response variable. The figure includes: (A) Predicted vs. Observed Values, showing the relationship between predicted and observed fexofenadine loads; (B) Residual Plot, displaying the residuals against predicted values to check for homoscedasticity; (C) Residuals Histogram, illustrating the distribution of residuals with a fitted kernel density estimate; (D) Normal QQ Plot, assessing the normality of residuals; (E) Scale-Location Plot, showing the spread of residuals to identify non-constant variance; and (F) Leverage Plot, indicating influential data points using Cook's distance.

Table SI 8: Ordinary Least Squares (OLS) regression results using airborne pollen concentrations of 14 allergenic taxa as predictors and baseline-subtracted fexofenadine loads as the response variable.

```
                                 OLS Regression Results
=======================================================================================
Dep. Variable:            fex_res_pos   R-squared (uncentered):                   0.899
Model:                            OLS   Adj. R-squared (uncentered):              0.893
Method:                 Least Squares   F-statistic:                              133.5
Date:                Fri, 31 May 2024   Prob (F-statistic):                    6.64e-96
Time:                        17:44:45   Log-Likelihood:                         -1443.4
No. Observations:                 223   AIC:                                      2915.
Df Residuals:                     209   BIC:                                      2963.
Df Model:                          14
Covariance Type:            nonrobust
==================================================================================
                     coef    std err          t      P>|t|      [0.025      0.975]
----------------------------------------------------------------------------------
Sweet_chestnut     3.1255      5.748      0.544      0.587      -8.206      14.457
Sorrel           -13.1799     12.033     -1.095      0.275     -36.901      10.541
Ragweed            4.6159     45.087      0.102      0.919     -84.268      93.500
Plantain          14.9983      5.157      2.908      0.004       4.832      25.165
Plane              5.6952      1.867      3.050      0.003       2.015       9.376
Oak                0.5800      0.994      0.584      0.560      -1.379       2.539
Mugwort         -151.1046     99.276     -1.522      0.130    -346.815      44.606
Hornbeam           3.1870      0.411      7.764      0.000       2.378       3.996
Hazel              0.1622      0.418      0.388      0.698      -0.661       0.986
Grasses            7.9020      0.318     24.877      0.000       7.276       8.528
Birch              1.4931      0.427      3.495      0.001       0.651       2.335
Beech              0.5571      0.645      0.864      0.389      -0.714       1.828
Ash                0.0594      0.422      0.141      0.888      -0.773       0.892
Alder              1.9006      0.596      3.188      0.002       0.725       3.076
==============================================================================
Omnibus:                       25.795   Durbin-Watson:                   1.009
Prob(Omnibus):                  0.000   Jarque-Bera (JB):               97.415
Skew:                           0.318   Prob(JB):                     7.02e-22
Kurtosis:                       6.175   Cond. No.                         590.
==============================================================================
```

**OLS with 19 allergenic pollen taxa**

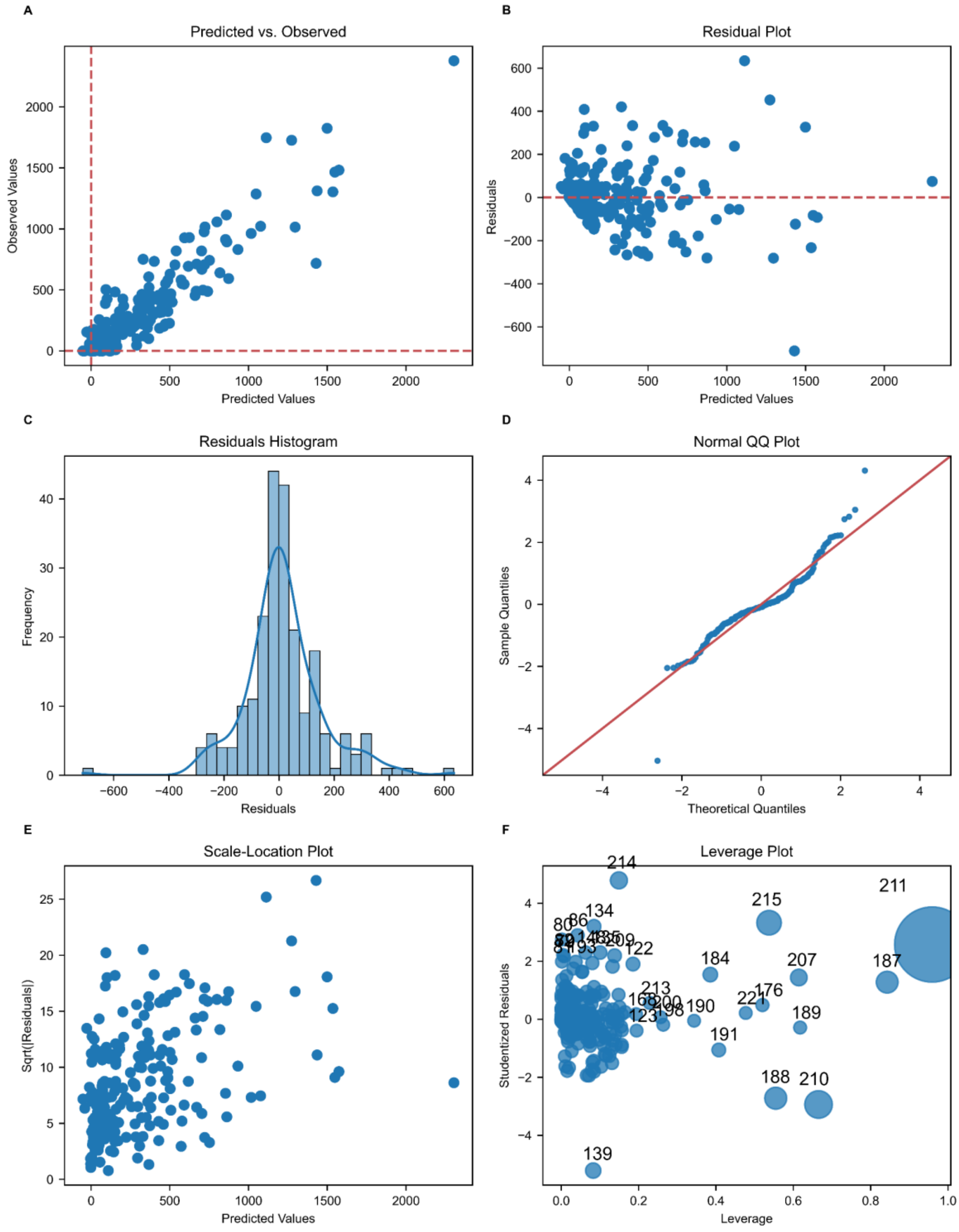


Figure SI 15: Diagnostic plots for the Ordinary Least Squares (OLS) regression analysis using airborne pollen concentrations of 19 allergenic taxa as predictors and baseline-subtracted fexofenadine loads as the response variable. The figure includes: (A) Predicted vs. Observed Values, showing the relationship between predicted and observed fexofenadine loads; (B) Residual Plot, displaying the residuals against predicted values to check for homoscedasticity; (C) Residuals Histogram, illustrating the distribution of residuals with a fitted kernel density estimate; (D) Normal QQ Plot, assessing the normality of residuals; (E) Scale-Location Plot, showing the spread of residuals to identify non-constant variance; and (F) Leverage Plot, indicating influential data points using Cook's distance.

Table SI 9: Ordinary Least Squares (OLS) regression results using airborne pollen concentrations of 19 allergenic taxa as predictors and baseline-subtracted fexofenadine loads as the response variable.

OLS Regression Results

| | | | |
|---|---|---|---|
| Dep. Variable: | fex_res_pos | R-squared (uncentered): | 0.914 |
| Model: | OLS | Adj. R-squared (uncentered): | 0.906 |
| Method: | Least Squares | F-statistic: | 114.5 |
| Date: | Fri, 31 May 2024 | Prob (F-statistic): | 1.08e-97 |
| Time: | 17:44:45 | Log-Likelihood: | -1425.7 |
| No. Observations: | 223 | AIC: | 2889. |
| Df Residuals: | 204 | BIC: | 2954. |
| Df Model: | 19 | | |
| Covariance Type: | nonrobust | | |

| | coef | std err | t | P>\|t\| | [0.025 | 0.975] |
|---|---|---|---|---|---|---|
| Yew | 0.1143 | 0.026 | 4.476 | 0.000 | 0.064 | 0.165 |
| Sweet_chestnut | 2.2295 | 5.411 | 0.412 | 0.681 | -8.440 | 12.899 |
| Sorrel | -4.3645 | 11.606 | -0.376 | 0.707 | -27.247 | 18.518 |
| Ragweed | 1.2135 | 42.159 | 0.029 | 0.977 | -81.910 | 84.337 |
| Plantain | 20.7211 | 5.131 | 4.039 | 0.000 | 10.605 | 30.837 |
| Plane | 5.9228 | 1.907 | 3.106 | 0.002 | 2.163 | 9.683 |
| Pine | -0.6388 | 0.241 | -2.655 | 0.009 | -1.113 | -0.164 |
| Olive_family | -0.6826 | 16.439 | -0.042 | 0.967 | -33.095 | 31.730 |
| Oak | 0.4177 | 0.946 | 0.441 | 0.659 | -1.448 | 2.284 |
| Nettle_family | -1.3041 | 0.403 | -3.239 | 0.001 | -2.098 | -0.510 |
| Mugwort | -66.1451 | 96.951 | -0.682 | 0.496 | -257.300 | 125.010 |
| Hornbeam | 3.1908 | 0.444 | 7.182 | 0.000 | 2.315 | 4.067 |
| Hazel | 0.9977 | 0.431 | 2.316 | 0.022 | 0.148 | 1.847 |
| Grasses | 8.4397 | 0.340 | 24.803 | 0.000 | 7.769 | 9.111 |
| Cypress_family | -0.0931 | 0.155 | -0.602 | 0.548 | -0.398 | 0.212 |
| Birch | 1.4958 | 0.402 | 3.719 | 0.000 | 0.703 | 2.289 |
| Beech | 0.3889 | 0.615 | 0.632 | 0.528 | -0.824 | 1.602 |
| Ash | 0.2259 | 0.566 | 0.399 | 0.690 | -0.891 | 1.343 |
| Alder | -1.2042 | 0.884 | -1.363 | 0.174 | -2.946 | 0.538 |

| | | | |
|---|---|---|---|
| Omnibus: | 29.539 | Durbin-Watson: | 1.112 |
| Prob(Omnibus): | 0.000 | Jarque-Bera (JB): | 166.105 |
| Skew: | 0.188 | Prob(JB): | 8.53e-37 |
| Kurtosis: | 7.211 | Cond. No. | 7.12e+03 |

Notes:
[1] $R^2$ is computed without centering (uncentered) since the model does not contain a constant.
[2] Standard Errors assume that the covariance matrix of the errors is correctly specified.
[3] The condition number is large, 7.12e+03. This might indicate that there are strong multicollinearity or other numerical problems.

**OLS with only significant pollen taxa**

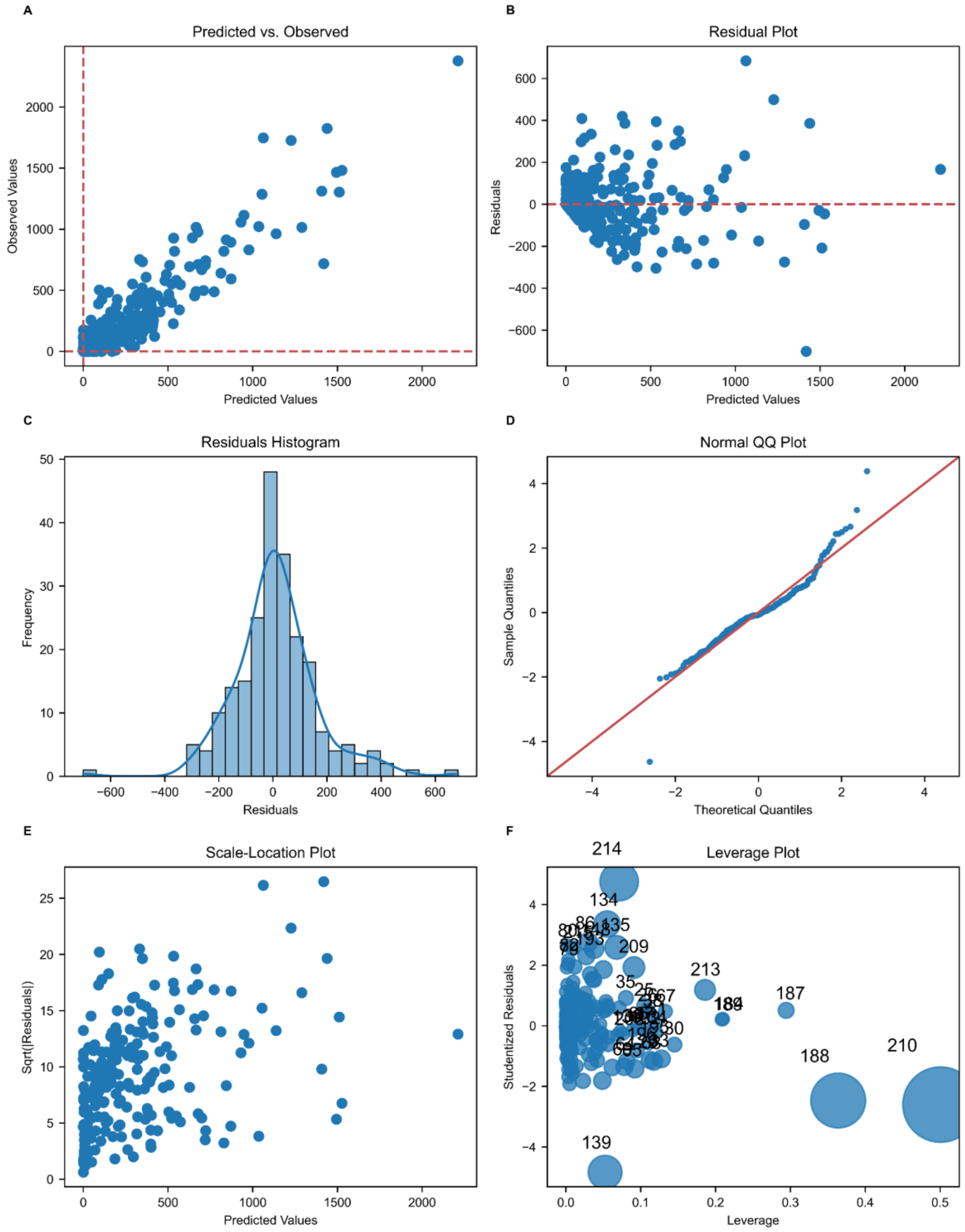


Figure SI 16: Diagnostic plots for the Ordinary Least Squares (OLS) regression analysis using airborne pollen concentrations of allergenic taxa with positive and significant coefficients ($p<0.01$) when considering 14 or 19 taxa as predictors, and baseline-subtracted fexofenadine loads as the response variable. The figure includes: (A) Predicted vs. Observed Values, showing the relationship between predicted and observed fexofenadine loads; (B) Residual Plot, displaying the residuals against predicted values to check for homoscedasticity; (C) Residuals Histogram, illustrating the distribution of residuals with a fitted kernel density estimate; (D) Normal QQ Plot, assessing the normality of residuals; (E) Scale-Location Plot, showing the spread of residuals to identify non-constant variance; and (F) Leverage Plot, indicating influential data points using Cook's distance.

Table SI 10: Ordinary Least Squares (OLS) regression results for allergenic taxa with positive and significant ($p < 0.01$) coefficients when considering 14 or 19 taxa as predictors, using baseline-subtracted fexofenadine loads as the response variable.

```
                                OLS Regression Results                                
=======================================================================================
Dep. Variable:            fex_res_pos   R-squared (uncentered):                   0.903
Model:                            OLS   Adj. R-squared (uncentered):              0.900
Method:                 Least Squares   F-statistic:                              286.5
Date:                Sat, 08 Jun 2024   Prob (F-statistic):                   1.43e-105
Time:                        12:15:20   Log-Likelihood:                         -1439.7
No. Observations:                 223   AIC:                                      2893.
Df Residuals:                     216   BIC:                                      2917.
Df Model:                           7                                                  
Covariance Type:            nonrobust                                                  
==============================================================================
                 coef    std err          t      P>|t|      [0.025      0.975]
------------------------------------------------------------------------------
Alder          0.4604      0.550      0.838      0.403      -0.623       1.544
Birch          1.6187      0.321      5.044      0.000       0.986       2.251
Grasses        7.7955      0.268     29.036      0.000       7.266       8.325
Hornbeam       3.1320      0.366      8.555      0.000       2.410       3.854
Plane          5.7669      1.702      3.389      0.001       2.413       9.121
Plantain      14.7647      3.745      3.942      0.000       7.383      22.147
Yew            0.0878      0.024      3.678      0.000       0.041       0.135
==============================================================================
Omnibus:                       28.364   Durbin-Watson:                   1.046
Prob(Omnibus):                  0.000   Jarque-Bera (JB):              115.445
Skew:                           0.351   Prob(JB):                     8.54e-26
Kurtosis:                       6.454   Cond. No.                         265.
==============================================================================

Notes:
[1] R² is computed without centering (uncentered) since the model does not contain a constant.
[2] Standard Errors assume that the covariance matrix of the errors is correctly specified.
```

#### NNLS and OLS model comparison

Table SI 11: Pollen-taxa specific weighting factors estimated using transformed concentrations of different combinations of pollen taxa as explanatory variables in NNLS and OLS models. Weighting factors in the OLS model with p-values below 0.01 are marked in bold.

Weighting factors (wp ; - )

| | Pollen taxa (explanatory variables) | 14 allergenic pollen taxa | | 19 allergenic pollen taxa | | All 47 pollen taxa and miscellaneous pollen |
|---|---|---|---|---|---|---|
| | Linear least squares regression type | NNLS | OLS | NNLS | OLS | NNLS |
| 1 | Alder | 1.902 | **1.901** | 0.000 | -1.204 | 0.000 |
| 2 | Amarant family | | | | | 0.000 |
| 3 | Ash | 0.004 | 0.059 | 0.008 | 0.226 | 0.000 |
| 4 | Beech | 0.621 | 0.557 | 0.614 | 0.389 | 0.206 |
| 5 | Birch | 1.562 | **1.493** | 1.570 | **1.496** | 0.768 |
| 6 | Buttercup family | | | | | 0.000 |
| 7 | Cedar | | | | | 0.000 |
| 8 | Common box | | | | | 0.000 |
| 9 | Crucifers | | | | | 0.000 |
| 10 | Cypress family | | | 0.000 | -0.093 | 0.000 |
| 11 | Daisy family | | | | | 0.000 |
| 12 | Elder | | | | | 0.000 |
| 13 | Elm | | | | | 4.429 |
| 14 | Fir | | | | | 0.000 |
| 15 | Ginkgo | | | | | 150.136 |
| 16 | Grasses | 7.813 | **7.902** | 7.858 | **8.440** | 7.861 |
| 17 | Hazel | 0.162 | 0.162 | 0.524 | 0.998 | 0.551 |
| 18 | Hophornbeam | NA | | | | 34.822 |
| 19 | Hornbeam | 3.194 | **3.187** | 3.184 | **3.191** | 3.191 |
| 20 | Horse chestnut | | | | | 0.000 |
| 21 | Larch | | | | | 0.000 |
| 22 | Lime-tree | | | | | 0.000 |
| 23 | Maize | | | | | 0.000 |
| 24 | Maple | | | | | 0.000 |
| 25 | Mercury | | | | | 0.000 |
| 26 | Miscellaneous | | | | | 0.000 |
| 27 | Mugwort | 0.000 | -151.105 | 0.000 | -66.145 | 0.000 |
| 28 | Mulberry family | | | | | 0.000 |
| 29 | Nettle family | | | 0.000 | **-1.304** | 0.000 |
| 30 | Oak | 0.154 | 0.580 | 0.142 | 0.418 | 0.000 |
| 31 | Olive family | | | 0.000 | -0.683 | 0.000 |
| 32 | Palm family | | | | | 47.880 |
| 33 | Pine | | | 0.000 | **-0.639** | 0.000 |
| 34 | Plane | 5.390 | **5.695** | 5.381 | **5.923** | 3.736 |
| 35 | Plantain | 11.433 | **14.998** | 11.785 | **20.721** | 8.265 |
| 36 | Poplar | | | | | 0.000 |
| 37 | Ragweed | 0.000 | 4.616 | 0.000 | 1.213 | 1.497 |
| 38 | Rose family | | | | | 0.000 |
| 39 | Rye | | | | | 1042.516 |
| 40 | Sedge family | | | | | 0.000 |
| 41 | Sorrel | 0.000 | -13.180 | 0.000 | -4.364 | 0.000 |
| 42 | Spruce | | | | | 0.000 |
| 43 | Sweet chestnut | 5.279 | 3.126 | 5.079 | 2.230 | 5.298 |
| 44 | Sweetgum | | | | | 46.420 |
| 45 | Umbellifers | | | | | 0.000 |
| 46 | Walnut | | | | | 0.000 |
| 47 | Willow | | | | | 5.879 |
| 48 | Yew | | | 0.087 | **0.114** | 0.079 |

#### Pollen multicollinearity

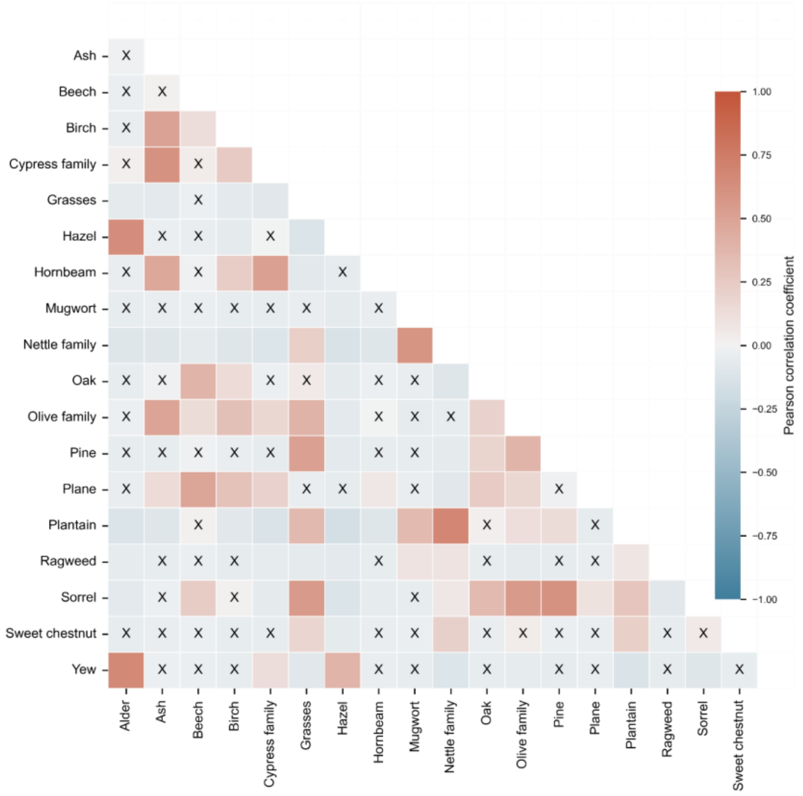


Figure SI 17: Correlation heatmap of airborne pollen concentrations from 19 allergenic pollen taxa. Pearson correlation coefficients, with values that are not significant (p > 0.05) marked with a cross.

#### NNLS and OLS timeseries contributions

##### NNLS 19 allergenic pollen taxa

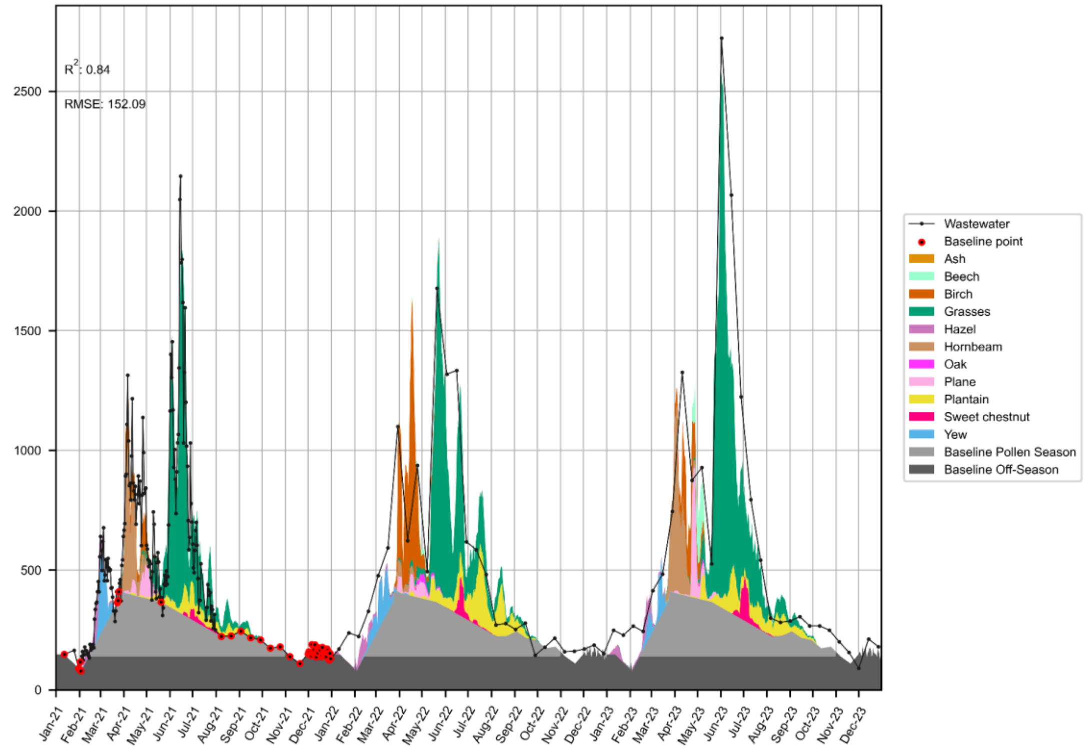


Figure SI 18: Observed fexofenadine wastewater loads in Zurich and daily contribution of single pollen taxa estimated from NNLS model, pollen season baseline, and off-season baseline. Pollen taxa-specific contributions to fexofenadine population loads estimated based on NNLS with concentrations of 19 allergenic pollen taxa and of total concentration of unidentifiable pollen (Miscellaneous) The colored areas represent the estimated pollen-taxa specific contributions calculated by multiplying the coefficient with the transformed pollen concentration of the respective pollen taxa and day. Baseline is determined by interpolation between fexofenadine loads on days in 2021 when no pollen taxa exceeded the lowest load class in the previous nine days. This baseline is further divided into baseline pollen season, which results from subtracting the baseline off-season (mean of fexofenadine loads in Jan., Dec., and Nov. 2021) values from the total baseline value of the respective day. The coefficient of determination (R2) and the root mean square error (RMSE) refer only to the prediction of the pollen-dependent fexofenadine loads.

**NNLS 47 pollen taxa**

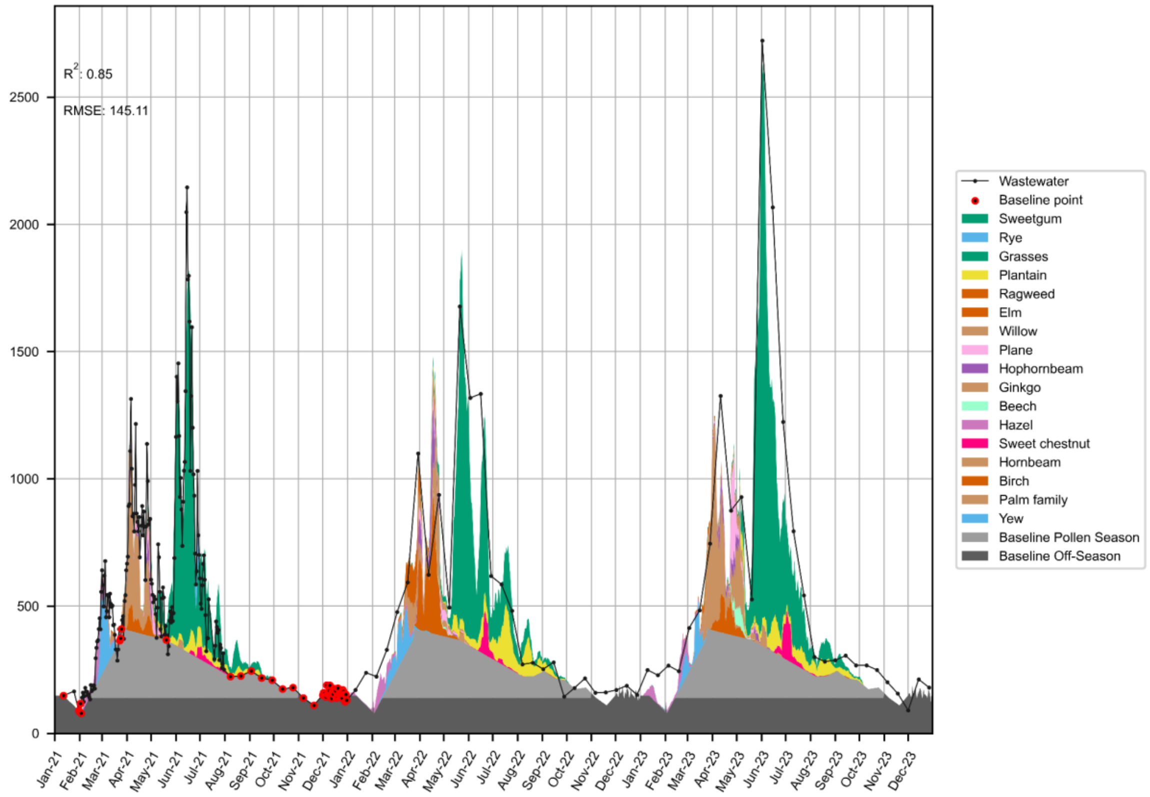


Figure SI 19: Observed fexofenadine wastewater loads in Zurich and daily contribution of single pollen taxa estimated from NNLS model, pollen season baseline, and off-season baseline. Pollen taxa-specific contributions to fexofenadine population loads estimated based on NNLS with concentrations of all 47 measured pollen taxa and of total concentration of unidentifiable pollen (Miscellaneous) The colored areas represent the estimated pollen-taxa specific contributions calculated by multiplying the coefficient with the transformed pollen concentration of the respective pollen taxa and day. Baseline is determined by interpolation between fexofenadine loads on days in 2021 when no pollen taxa exceeded the lowest load class in the previous nine days. This baseline is further divided into baseline pollen season, which results from subtracting the baseline off-season (mean of fexofenadine loads in Jan., Dec., and Nov. 2021) values from the total baseline value of the respective day. The coefficient of determination (R2) and the root mean square error (RMSE) refer only to the prediction of the pollen-dependent fexofenadine loads.

**OLS significant pollen taxa coefficients**

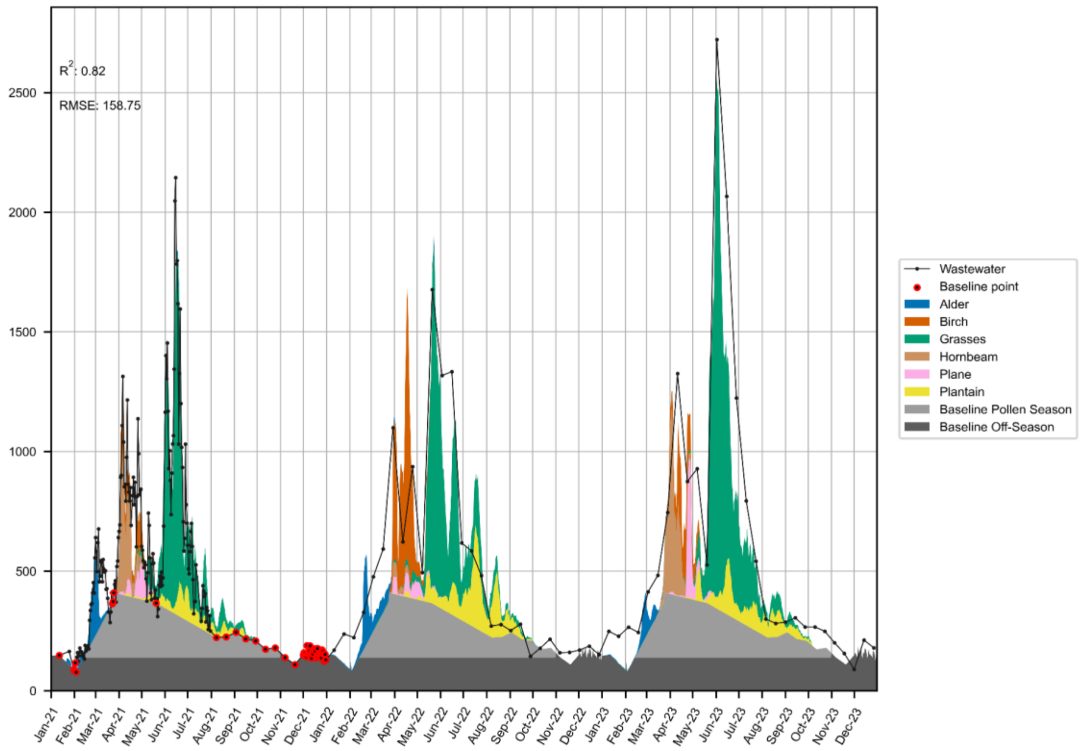


Figure SI 20: Observed fexofenadine wastewater loads in Zurich and daily contribution of single pollen taxa estimated from OLS model, pollen season baseline, and off-season baseline. Pollen taxa-specific contributions to fexofenadine population loads estimated based on OLS with concentrations of all 6 measured pollen taxa identified as significant (p<0.01) contributors in OLS with 14 allergenic taxa. The colored areas represent the estimated pollen-taxa specific contributions calculated by multiplying the coefficient with the transformed pollen concentration of the respective pollen taxa and day. Baseline is determined by interpolation between fexofenadine loads on days in 2021 when no pollen taxa exceeded the lowest load class in the previous nine days. This baseline is further divided into baseline pollen season, which results from subtracting the baseline off-season (mean of fexofenadine loads in Jan., Dec., and Nov. 2021) values from the total baseline value of the respective day. The coefficient of determination (R2) and the root mean square error (RMSE) refer only to the prediction of the pollen-dependent fexofenadine loads.

**Modeled contributions to annual fexofenadine wastewater loads**

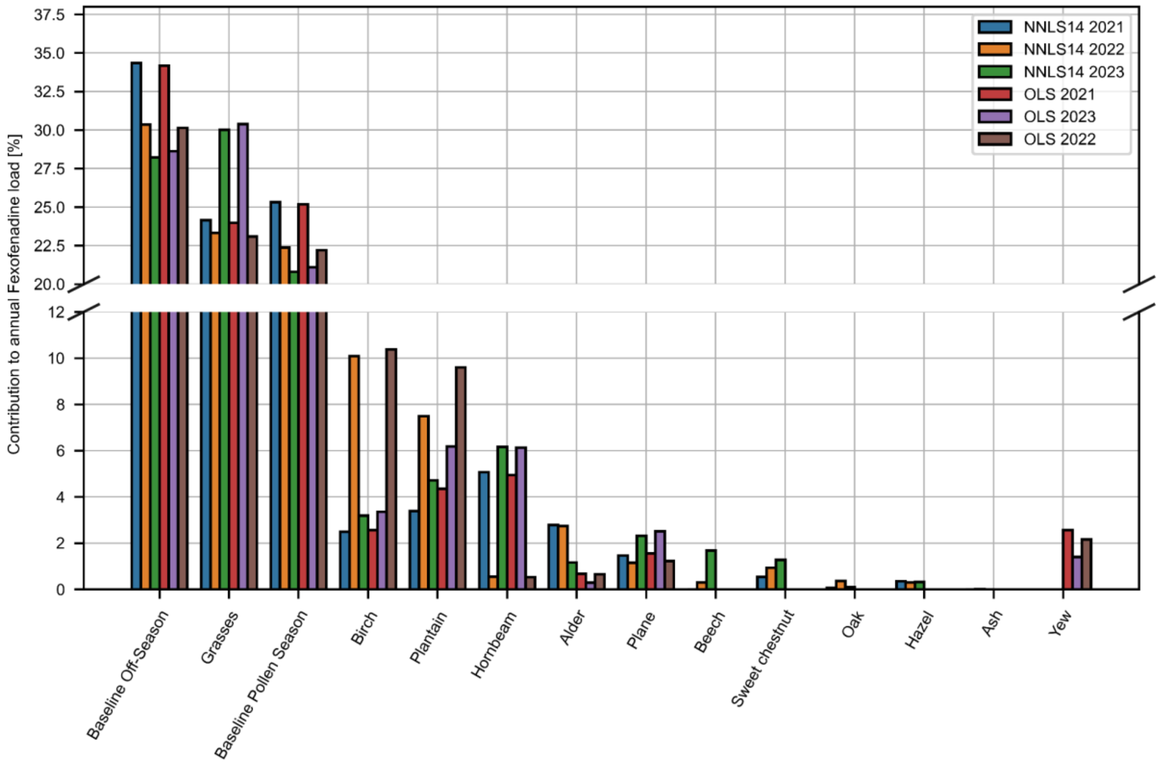


Figure SI 21: Relative contributions of baseline and specific pollen taxa to annual fexofenadine wastewater loads. The results are presented for two models: one considering 14 allergenic pollen taxa in NNLS (NNLS14), and the OLS model, in which only the pollen taxa with positive and significant (p<0.01) coefficients were considered.

### SI 3. In-sample stability of antihistamines

#### SI 3.1 Sample preparation and analytical procedure

The stability of the target analytes in both spiked wastewater and Evian mineral water was examined at different storage temperatures (-20°C, 4°C, and 22°C).

As no pharmaceutical products containing rupatadine are licensed in Switzerland and the compound was not detected in wastewater, rupatadine was excluded from the stability experiments.

For the wastewater matrix, a 24-hour composite influent sample was collected under dry weather conditions from the Zurich Werdhölzli WWTP (28.03.2022). The wastewater sample was stored in a muffled 1000ml Schott DURAN borosilicate glass bottle from Kisker Biotech and stored at 4°C in the dark for four days until the start of the experiment. The bottle was then gently shaken and left for 1.5h to allow larger particles to settle. 700mL of the supernatant was then transferred to a muffled bulkhead bottle and spiked with an ethanol-based substance mix (1.7mL) to a final concentration of 1000ng/L. Cetirizine was spiked independently from an acetonitrile-water (1:1, v/v) based solution to a final concentration of 4000ng/L. The bottle was then shaken and left stand for equilibration for 2h. 20mL were taken out to measure the pH (SevenMulti, Mettler Toledo) before aliquoting the sample into muffled 1.5mL MS glass vials (BGB Analytik). The same procedure was used for the preparation of the Evian mineral water samples.

At the beginning of the experiment, the aliquots were continuously incubated at their corresponding temperature condition. After the incubation period had elapsed, the respective sample was thawed for 1 minute under lukewarm water (only for frozen samples), spiked with 20 µL of a 60 µg/L ethanol-based isotope-labelled internal standard solution, shaken and vortexed, centrifuged at room temperature for 10 min at 279.5 RCF (5427 R, Eppendorf centrifuge) and measured immediately. All samples were analyzed by large volume direct injection reversed-phase LC-HRMS, as described in the methods section.

Samples of the same matrix were treated as replicates for the first measurement ($t_0$). At most time points only one sample condition was measured but occasionally replicate measurements were performed as described in the table below.

Table SI 12: Experimental conditions of in-sample stability experiment and number of samples measured per condition and time point

| | Wastewater Influent | | | Evian mineral water | |
|---|---|---|---|---|---|
| Time point | 22°C | 4°C | -20°C | °4C | -20°C |
| 0h | 3 | 3 | 3 | 3 | 3 |
| 1d | 1 | 1 | 1 | 1 | 1 |
| 2d | 1 | 1 | 1 | 1 | 1 |
| 3d | 1 | 1 | 1 | 1 | 1 |
| 7d / 8d | 2 | 2 | 2 | 2 | 2 |
| 9d / 10d | 2 | 2 | 2 | 2 | 2 |
| 14d / 15d | 2 | 2 | 2 | 2 | 2 |
| 20d | 1 | 1 | 1 | 1 | 1 |
| up to 286d | - | - | 1 | - | 1 |

Table SI 13: Result of pH measurements of the stability experiment samples after spiking and at the end of experiment

| Matrix | Temperature [°C] | pH | |
|---|---|---|---|
| | | After spiking | End of experiment (t=20d) |
| Wastewater influent | 22 | 7.6 | 7.6 |
| | 4 | 7.6 | 5.9 |
| | -20 | 7.6 | 8.2 |
| Evian mineral water | 4 | 7.7 | 8.2 |
| | -20 | 7.7 | 9.5 |

#### SI 3.2 Data analysis

The change in relative concentration over time was assessed by comparing the peak area ratio of the analyte to its corresponding ISTD. The average response ratio of the triplicate at $t_0$ was set to 100% for each substance and condition. The change in relative concentration was then expressed as share of the initial area ratio to determine degradation or formation over time.
A non-linear pseudo first order kinetic model was fitted to the data to determine the degradation rate constant for each substance in the specific matrix and at the particular condition. The degradation function is described in Equation SI 7, where $A_0$ represents the initial response ratio, *A* equals the response ratio at a particular measurement time point, *k* ($d^{-1}$) stands for the degradation rate constant and t for a point in time (d)

$$A = A_0 e^{-kt} \qquad (SI\,7)$$

The parameters *A0* and *k* were optimized using Scipy in Python.

#### SI 3.3 Stability results

Fexofenadine, cetirizine, and diphenhydramine, when stored at 22°C for 20 days or at -20°C for 280 days, demonstrated consistent stability in wastewater, maintaining relative concentrations within the 80% to 120% range. In contrast, bilastine remained stable at 22°C for 20 days but exhibited accelerated degradation at -20°C, dropping to approximately 60% of its initial concentration after 280 days (Figure SI 22 and Figure SI 23).

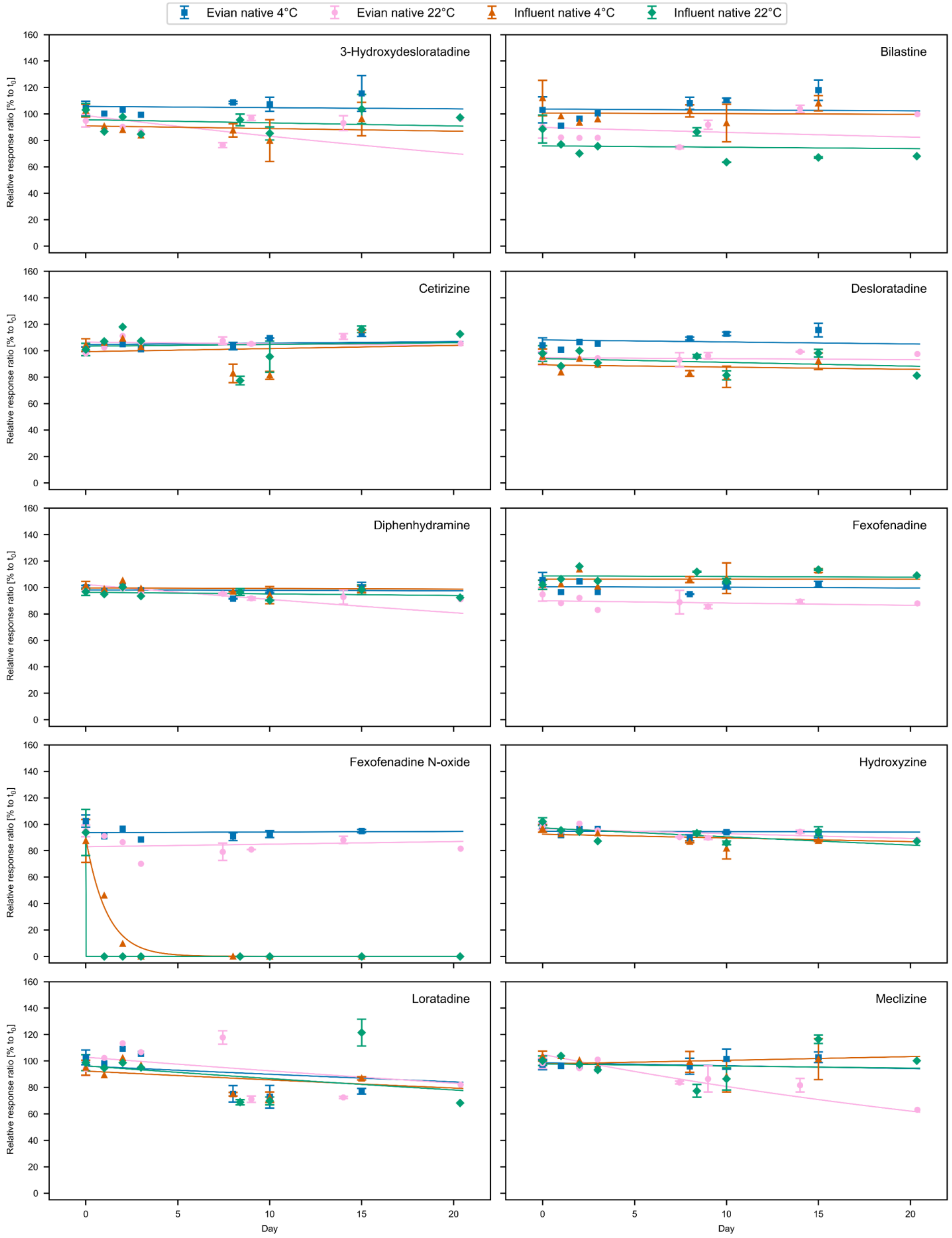


Figure SI 22: In-sample stability of antihistamines in wastewater influent and Evian mineral water incubated at 4°C and 22°C over a period of 20 days. Measured data (points) and pseudo first order kinetics fit (lines) are displayed.

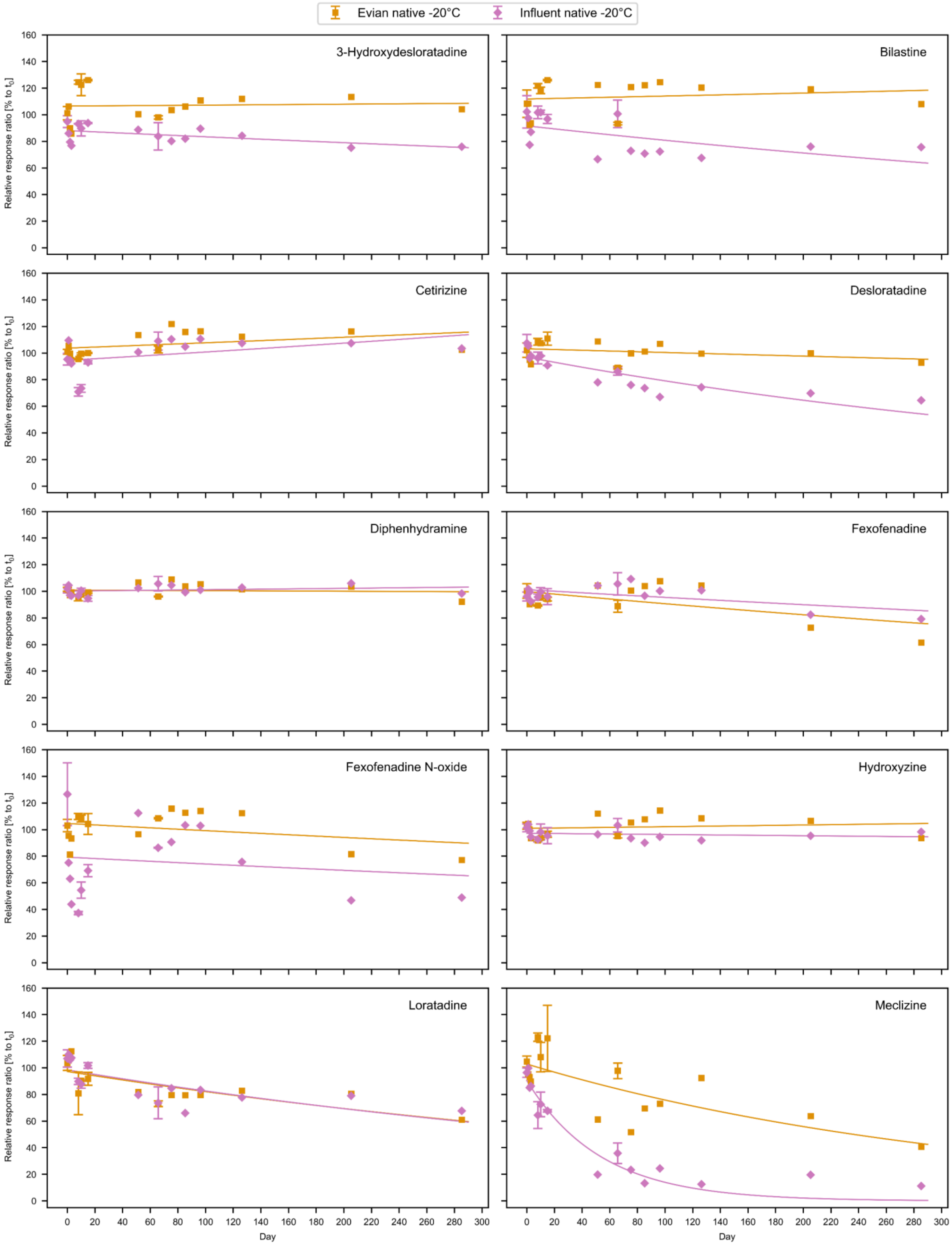


Figure SI 23: In-sample long-term stability of antihistamines in wastewater influent and Evian mineral water incubated at -20°C. Measured data (points) and pseudo first order kinetics fit (lines) are displayed.

#### SI 4. References


1. Gehrig Bichsel R, Maurer F, Schw ierz C. *Regionale Pollenkalender Der Schweiz. Fachbericht MeteoSchweiz, 264.*; 2017.
